# Multiple and novel molecular mechanisms in *TUBA1A*-related tubulinopathy: insights from deep clinical and neuroradiological phenotyping

**DOI:** 10.1101/2025.03.28.25324751

**Authors:** James Fasham, Rucha Pandit, Adam T Higgins, Dowon Kim, Karl Austin-Muttitt, Anna V Derrick, Robert Gunn, Maya Matty, Nicola Ragge, Kate E Chandler, Gabriela E Jones, Mohnish Suri, Alison Kraus, Tabib Dabir, Katrina Stone, Wendy D Jones, Julia Rankin, James A Poulter, Marilena Elpidorou, Eamonn Sheridan, Colin A. Johnson, Helen V Firth, Siddharth Banka, Ruth Newbury-Ecob, Neeti Ghali, Andrew J Green, Shelagh Joss, Sahar Mansour, Pradeep Vasudevan, Michael J Parker, Jonathan GL Mullins, Adam G Thomas, Seo-Kyung Chung, Abhijit Dixit

**Affiliations:** Department of Clinical and Biomedical Sciences, Faculty of Health and Life Sciences, University of Exeter, Exeter, EX2 5DW, UK; Peninsula Clinical Genetics Service, Royal Devon & Exeter Hospital (Heavitree), Gladstone Road, Exeter, EX1 2ED, UK; Kids Neuroscience Centre, Kids Research, Children’s Hospital at Westmead, Sydney, NSW 2145, Australia; Swansea University Medical School, Swansea University, Swansea, SA2 8PP, UK; Brain and Mind Centre, Faculty of Medicine and Health, University of Sydney, NSW 2050, Australia; West Midlands Regional Genetics Service, Birmingham Women and Children’s NHS Foundation Trust, Birmingham B15 2TG, UK; Faculty of Health and Life Sciences, Oxford Brookes University, Oxford OX3 0FL, UK; Manchester Centre for Genomic Medicine, 6th Floor St Mary’s Hospital, Oxford Road, Health Innovation Manchester, Manchester, M13 9WL, UK; Department of Clinical Genetics, Nottingham University Hospitals NHS Trust, City Hospital Campus, Hucknall Road, Nottingham NG5 1PB, UK; Yorkshire Regional Genetics Service, Chapel Allerton Hospital, Chapeltown Rd, Leeds LS7 4SA, UK; Northern Ireland Regional Genetics Service, Belfast City Hospital. Lisburn Rd. Belfast. Northern Ireland BT0 8AB, UK; North East Thames Regional Genetics Service, Great Ormond Street Hospital for Children NHS Foundation, Guilford St, London WC1N 3BH, UK; Leeds Institute of Medical Research, University of Leeds, St. James’s University Hospital, Beckett St, Harehills, Leeds LS9 7TF, UK; Clinical Genetics, Box: 134, Level 6, Addenbrooke’s Treatment Centre, Cambridge University Hospitals NHS Foundation Trust, Hills Road, Cambridge, CB2 0QQ, UK; Division of Evolution, Infection and Genomics, School of Biological Sciences, Faculty of Biology, Medicine and Health, University of Manchester, Manchester Academic Health Science Centre, Manchester M13 9PT, UK; Department of Clinical Genetics, Level B, St Michael’s Hospital, Bristol, BS2 8EG, UK; North West Thames Regional Genetics Service, London North West Healthcare University NHS Trust, Northwick Park Hospital, A404 Watford Rd, Harrow, Middlesex, HA1 3UJ, UK; Department of Clinical Genetics, Our Lady’s Children’s Hospital, Crumlin, Dublin, D12 N512, Ireland; Department of Clinical Genetics, Queen Elizabeth University Hospital, 1345 Govan Rd, Glasgow G51 4TF, UK; Molecular and Clinical Sciences Research Institute, St. George’s University of London, Cranmer Terrace, London SW17 0RE, UK; South West Thames Regional Genetics Service, St. George’s Hospital, Blackshaw Rd, London SW17 0QT, UK; Department of Clinical Genetics, University Hospitals of Leicester NHS Trust, Leicester LE1 5WW, UK; Sheffield Clinical Genetics Service, Sheffield Children’s NHS Foundation Trust, Northern General Hospital, Herries Road, Sheffield S5 7AU, UK; Department of Neuroradiology, Queens Medical Centre, Nottingham University Hospitals NHS Trust, Derby Road, Nottingham, NG7 2UH, UK

**Keywords:** Tubulin, lissencephaly, malformation of cortical development, MCD, penetrance, expressivity

## Abstract

**Purpose:** Heterozygous *TUBA1A* variants are a well-recognised cause of malformations of cortical development (MCDs). Although existing literature − predominantly radiologically ascertained cohorts − suggests complete penetrance of the MCD phenotype in this condition, there is also anecdotal contrary evidence. Understanding the clinical spectrum of *TUBA1A*-related disorders informs counselling and clinical decisions, especially for those identified early in life.

**Methods:** Individuals with *TUBA1A* variants were identified through a large exome sequencing project (DDD project) and analogous studies. Additional clinical data and independent review of the neuroradiology were obtained. Functional studies of the incorporation, reincorporation and depolymerisation of variant tubulin molecules, were performed by expressing wild-type and variant TUBA1A in HEK293 cells.

**Results:** We identified 23 individuals with 20 pathogenic/likely pathogenic *TUBA1A* variants (10 novel). Nearly all had characteristic neuroradiological features of tubulinopathies, but only 25% had MCDs (vs. 99% in previous studies). The variants identified in this study are distinct from those previously described, potentially highlighting a genotype-phenotype relationship, and have a variety of effects on microtubule formation, including reduced depolymerisation that has not been previously observed in *TUBA1A*-related tubulinopathies.

**Conclusion:** Our findings support a lower penetrance of MCDs in *TUBA1A*-related tubulinopathy, with immediate relevance to variant interpretation and genetic counselling for this condition.

## Introduction

Malformations of cortical development (MCD) are a large group of neurodevelopmental disorders that encompass neuronal migration disorders including lissencephaly and polymicrogyria.^1^ Clinically, these usually result in severe developmental impairment and seizures, significantly impacting quality of life.^2^ Neuronal migration is a complex process by which neurons travel radially from their point of origin in the central developing brain to form complex, often laminar, structures. Abnormalities affecting the tubulin proteins, termed tubulinopathies, and specifically variants in the gene *TUBA1A* impact neuronal migration and are a well-recognised genetic cause of MCDs.^3^ Indeed, for certain phenotypic subgroups, such as lissencephaly with cerebellar hypoplasia, heterozygous variants in *TUBA1A* account for 30% of cases.^4^

*TUBA1A* encodes a tubulin alpha subunit, a globular protein that forms heterodimers with a number of tubulin beta subunits. These alpha-beta tubulin dimers in-turn polymerise to form microtubules.^5^ Microtubes are long hollow cylindrical polymers that are pivotal to a number of cellular functions including defining and maintaining cytoskeletal structure and guiding cell division, intracellular transport and neuronal migration.^6^ Alpha tubulins are ubiquitous in eukaryotes; consequently there are a large number of orthologous proteins to *TUBA1A*. There are also a number of human paralogues that show differential temporal and tissue expression at different life-stages. Throughout these numerous homologues, - the domain architecture of the protein, including functionally important sites such as the invariant N-terminal domain, a non-exchangeable GTP binding site, plus binding sites for the molecular motors kinesins and dyneins, and the C-terminal amino acid sequence are remarkably conserved.^7^ *TUBA1A*, in common with other dominant tubulinopathy genes, has a very high missense constraint score (8.75 on gnomAD v4.1.0), suggesting exquisite intolerance to missense variation.

The first human disorder associated with pathogenic variants in *TUBA1A* was described in 2007.^8,9^ Eighteen years later, there are over 100 published individuals with *TUBA1A*-related tubulinopathy. The vast majority of these have either radiographic or neuropathological evidence of an MCD,^10^ although case reports, individuals in diagnostic exome studies,^11,12^ and unpublished clinical reports have raised uncertainty regarding such high penetrance. Since the ascertainment of all large case series published to date has been based on the evidence of either radiographic or histological cortical malformation, they are unable to adequately address this issue. This study, using an alternative strategy for case identification involving a national project with broad recruitment criteria (the DDD project),^13^ aims to address the penetrance question; *In vitro* functional validation is also presented to add robust evidence for pathogenicity and illuminate functional differences between the identified *TUBA1A* variants.

## Materials and Methods

### Patient ascertainment, clinical and genetic studies

A Complementary Analysis Project (CAP) permitted the authors to identify all probands with proven or presumed *de novo TUBA1A* variants recruited to the Deciphering Developmental Disorders (DDD) project. This was a research project that recruited children with developmental disorders through 24 UK and Irish regional Clinical Genetics centres between 2011 and 2015. Each patient, having usually already undergone standard-of-care diagnostic genetic testing including single gene or panel sequencing, underwent microarray and exome sequencing. The latter was typically performed as a trio, with both parents recruited alongside the proband. An automated variant filtering pipeline was then applied to prioritise candidate variants. The sequencing and filtering methodologies have previously been described in full.^14^

Clinical data was extracted from the Decipher platform (www.deciphergenomics.org) and/or obtained directly from the responsible clinician using a proforma. Microcephaly was defined as a head circumference greater than two standard deviations below the mean as per the human phenotype ontology (HPO) definition (https://hpo.jax.org) to allow comparison with Hebebrand *et al.* 2019.

### Neuroradiology

Brain magnetic resonance imaging (MRI), performed as part of participants’ routine clinical care, was reviewed by a single expert reviewer (A.T.). Each neuroradiological feature (cortex, basal ganglia, corpus callosum, cerebellum, brainstem) was graded (0, normal; 1, mild changes; 2, moderate changes; 3, severe changes). The anterior commissure was also graded (0, normal; 1, hypoplastic; 3, absent). Scores were summated to approximate an overall radiological severity score out of maximum 18. These were divided into mild (0-6), moderate (7-12) and severe (13-18) categories. Non-visualization of the internal capsule on axial plane images through the basal ganglia has been described as a characteristic feature of tubulinopathies. However, review of coronal imaging demonstrates that the capsule is normally present but rotated downward into the axial plane. In order to quantify the extent of rotation, the interhemispheric falx was taken as a reference point for the vertical and the angle subtended by a second line was drawn through the anterior limb of the internal capsule in the coronal plane at the level of the hypopthalamus/mamillary bodies was recorded on each side.

### In silico modelling

Protein structural models for TUBA1A wild-type and variants were predicted using a homology modelling pipeline (Mullins, 2012). A previously published template derived from electron microscopy (EM) data was used to model microtubule structure (PDB: 2XRP).^15^ UCSF Chimera package was used to visualise and examine the *in silico* protein models (www.cgl.ucsf.edu/chimera).

### Functional studies

#### Expression construct generation

Wild-type TUBA1A was expressed in a C-terminally DDK-tagged expression construct (pRK5). The QuickChange II Site-Directed Mutagenesis kit (Agilent) was used to introduce variants into the wild-type (WT) expression construct. Both WT and variant constructs were transformed into chemically competent *E. coli* cells (NEB) using the heat-shock method according to the manufacturer’s instructions and were sequence-validated following miniprep plasmid isolation (Qiagen).

#### Cell culture and transfection

Human embryonic kidney (HEK293) cells were cultured and maintained as previously described.^16^ 8×10^5^ cells were seeded on 6 cm dishes (Thermo Fisher Scientific) for protein extraction and western blotting. 4×10^4^ cells were seeded on poly-D-lysine (Sigma-Aldrich) coated glass coverslips (16mm) for immunofluorescence imaging. 24 hours post seeding, the cells were transfected with DDK-tagged wild-type and variant expression constructs using TurboFectin 8.0 (OriGene).

#### Immunofluorescence

24 hours post transfection, HEK293 cells were fixed in methanol at −20°C for 3 minutes and permeabilised with 0.5% Triton X-100 (Sigma-Aldrich) in phosphate-buffered saline (PBS; Thermo Fisher Scientific) to facilitate the imaging of intracellular tubulin epitopes. Fixed cells were blocked with PBS containing 0.1% Triton X-100 and 2% w/v Bovine Serum Albumin (BSA). Primary antibodies for staining included anti-DDK (mouse monoclonal IgG2a; 1:1500; OriGene) and anti-α-tubulin (rabbit polyclonal; 1:1500; Abcam) followed by goat anti-mouse Alexa Fluor 568 and goat anti-rabbit Alexa Fluor 488 (1:600; Invitrogen). Hoechst (1:10000; Thermo Fisher Scientific) was used as a cell nucleus marker. ProLong Gold Antifade Mountant (Thermo Fisher Scientific) was used to mount the immunostained coverslips onto glass microscope slides. Images (magnification 63X) were acquired on a confocal microscope (Zeiss LSM 710) and analysed using the ImageJ software (NIH).

#### Protein extraction and western blotting

24 hours post-transfection, HEK293 cells were lysed and sequentially extracted into the cytosolic and microtubule fractions. Extractions were performed at 37°C for physiological incorporation (C1) and at 4°C for depolymerisation experiments. Briefly, the cells were lysed in the microtubule stabilisation buffer containing 80mM PIPES buffer, 1mM of MgCl_2_ and EGTA, 10% glycerol, 0.5% Triton-X, 0.001% Antifoam and 1% protease inhibitor cocktail (PIC; Sigma-Aldrich). This yields the cytosolic fraction, which was precipitated using ice-cold acetone (Sigma-Aldrich) and the precipitate was dissolved in M-PER (Thermo Fisher Scientific) containing 1% PIC. The pellet from cell lysis was resuspended in the microtubule destabilisation buffer containing 20mM Tris HCl, 150mM NaCl, 10 mM CaCl_2_, 1% Triton-X and 1% PIC and centrifuged to yield proteins in the microtubules as the supernatant (insoluble fraction). Proteins in the cytosolic and microtubule fractions were quantified using the DC protein assay (Bio-Rad).

For the depolymerisation experiments, 24 hours post-transfection, transfected HEK293 cells were incubated at 4°C for different timepoints (2min, 5min and 10min) before extracting the cytosolic and microtubule fractions as described above. For the repolymerisation experiments, transfected HEK293 cells were incubated at 37°C for 15min after incubating at 4°C for 30min.

Cell proteins from both fractions were separated on a 4-12% Bis-Tris gel (Bolt, Thermo Fisher Scientific) with MES SDS running buffer (Thermo Fisher Scientific). Separated proteins were transferred on a 0.45µm nitrocellulose membrane using the Turbo TransBlot system (semi-dry transfer; Bio-Rad) and total protein stain (Revert; LI-COR) was used as a loading control following manufacturer’s instructions. Membranes were blocked with Odyssey blocking buffer (LI-COR) and Tri-buffered saline (TBS) and immunoblotted with primary anti-DDK (IgG2a) antibody (mouse; 1:3000; OriGene) and anti-β-actin (rabbit; 1:5000; Abcam) as an additional housekeeping control. The proteins were visualised using secondary antibodies 800CW anti-mouse (1:25,000; LI-COR) and 680RD anti-rabbit (1:20,000; LI-COR) on the Odyssey imaging system (LI-COR) and quantified using Image Studio Software (LI-COR).

#### Statistical Analysis

Absolute ratio of wild-type insoluble fraction was standardised to 100% for each blot and was used to calculate the percent insoluble ratio for the variants. Statistical analyses were performed on Prism9 (GraphPad) using a one-way ANOVA with multiple comparisons (Dunnett’s test). All values were expressed as mean ± SEM, unless otherwise stated.

## Results

### Cohort composition

Nineteen individuals with likely causative, proven or presumed *de novo* variants in *TUBA1A* were identified from 13,600 patients recruited to the DDD project (**Table 1**). 1/19 declined invitation and the identical twin of one DDD proband (**Individual 4**) was added. Four further individuals were identified, all with presumed causative *TUBA1A* variants identified through gene-agnostic sequencing strategies analogous to those utilised in the DDD study. For one individual (**Individual 14**) a presumed *de novo TUBA1A* variant was identified by genome sequencing performed as part of the Genomics England 100,000 Genomes Project^17^ and for three individuals *TUBA1A* variants were identified by either diagnostic exome (**Individual 12**, **Individual 19**) or diagnostic genome (**Individual 5**) sequencing performed as part of clinical care (for methods see **Supplemental text**). Thus, a study cohort of 23 individuals was assembled.

**Table 1:**
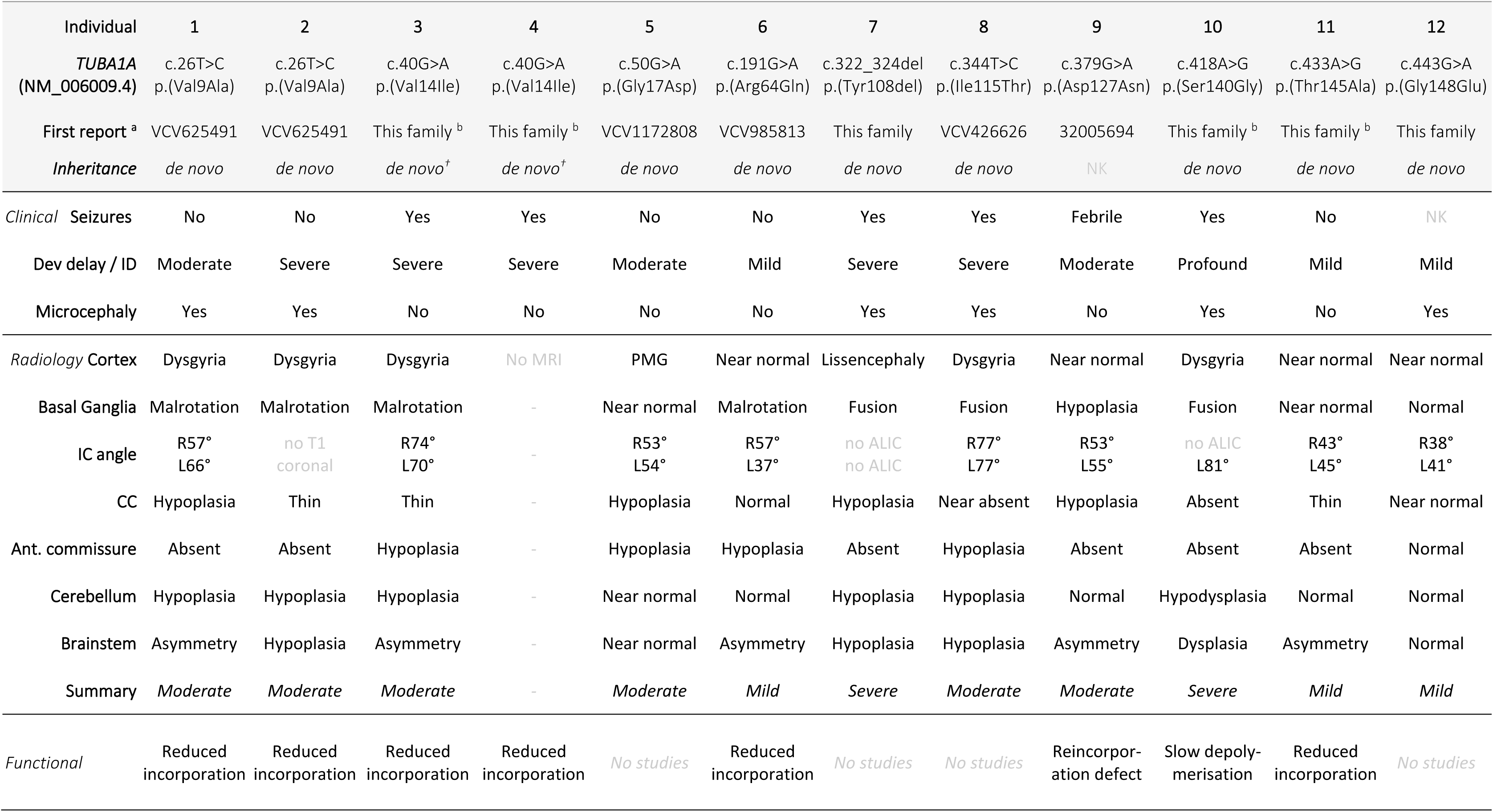
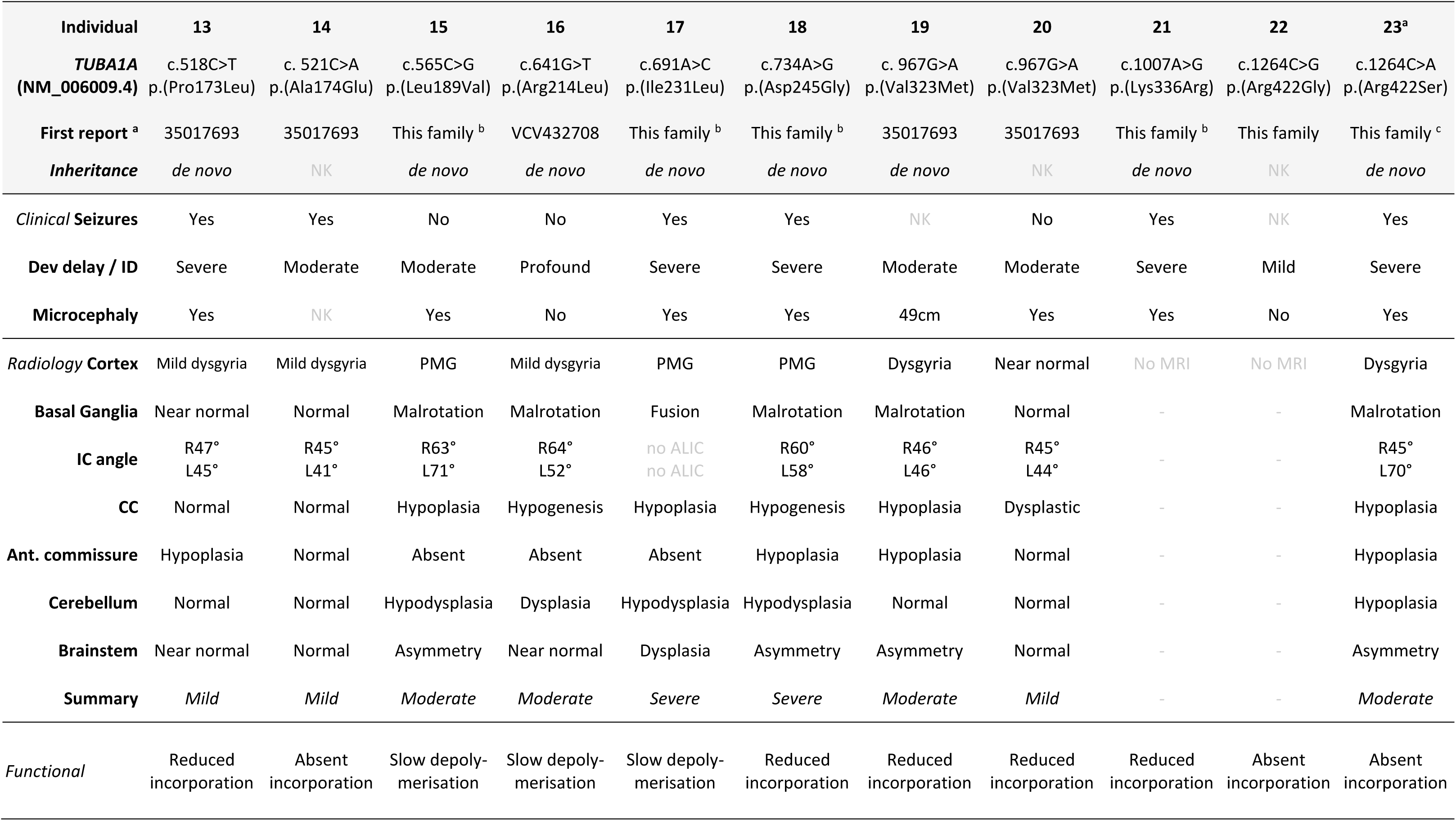
Clinical features of individuals with pathogenic *TUBA1A* variants. ^a^ Published articles these are shown in preference to ClinVar entries regardless of chronology. ^b^ The variants are referenced in a review^10^ that drew summary data from the DDD study, but did not publish individual-level clinical data. It is extremely likely, but not certain, that the individual referred to in this study are the individuals in our study. ^c^ The clinical details of this individual have since been published.^12^ *Abbreviations:* †, individuals are identical twins; Abn. Abnormal; ALIC Anterior limb of the internal capsule; CC, corpus callosum; Dev. Delay, Developmental delay; IC, internal capsule; ID, intellectual disability; L, left; MRI, magnetic resonance imaging; NK, not known; PMG, Polymicrogyria; R, right.

### *TUBA1A* variants

Twenty unique proven or presumed *de novo* heterozygous *TUBA1A* variants were identified. Three of these were present in two individuals **(Supplemental Table 1)**. The identified variants are shown alongside population allele frequencies and predictions from *in silico* missense prediction tools. 19/20 *TUBA1A* variants were missense substitutions and one was an in-frame deletion of a single amino acid, NM_006009.4: c.322_324del p.(Tyr108del). The variants identified were completely absent from population databases, with the exception of p.(Arg64Gln), which is present in one individual in UKBiobank and hence gnomAD 4.1.0. The individual in UKBiobank with the p.(Arg64Gln) has a phenotype compatible with a tubulinopathy, although neuroradiology was not available to review. The majority of missense variants were predicted damaging by *in silico* pathogenicity meta prediction tools REVEL (>0.7) and AlphaMissense. All were classified as either pathogenic or likely pathogenic according to the ACMG/AMP criteria^18^ (**Supplemental Table 2**) and in no case was another more likely causative variant identified (**Supplemental Table 3**). Variants were predominant present in, and evenly distributed throughout, the N-terminal half of the TUBA1A protein, mostly comprising the GTPase domain (**Supplemental Figure 1**).

Clustering of pathogenic *TUBA1A* variants in the 3’ region of the protein,^10,12^ apparent in studies that have employed neuroradiological or pathological ascertainment strategies, was not observed in our study. Further, p.(Arg264Cys) and p.(Arg402His), that account for an estimated 13% of previously reported affected individuals,^10^ were not observed. Novel recurrent variants were observed in this cohort, including NM_006009.4: c.40G>A; p.(Val14Ile), shared by identical twins and NM_006009.4: c.26T>C; p.(Val9Ala) and NM_006009.4: c. 967G>A; p.(Val323Met), each occurring in two unrelated individuals. For 18/20 variants, our cohort represents the first descriptions of associated human disease, although in one case this has since been published,^12^ and a prior study has listed some variants from our families without their individual-level clinical phenotypes.^10^

### Neuroradiological findings

Neuroimaging was available for 20/23 individuals (**Table 1**): **Individual 4** did not have neuroimaging although this was available for their identical sibling; **Individual 21** died before this study commenced without neuroimaging being performed; **Individual 22** is an adult with intellectual impairment for whom neuroimaging was not clinically indicated. Exemplar findings are shown in **Figure 1**. Individual participant images and reports (**Supplementary Text**, **Supplemental Table 4**) with neuroradiological scores (**Supplemental Table 5**) are also included.

**Figure 1:**
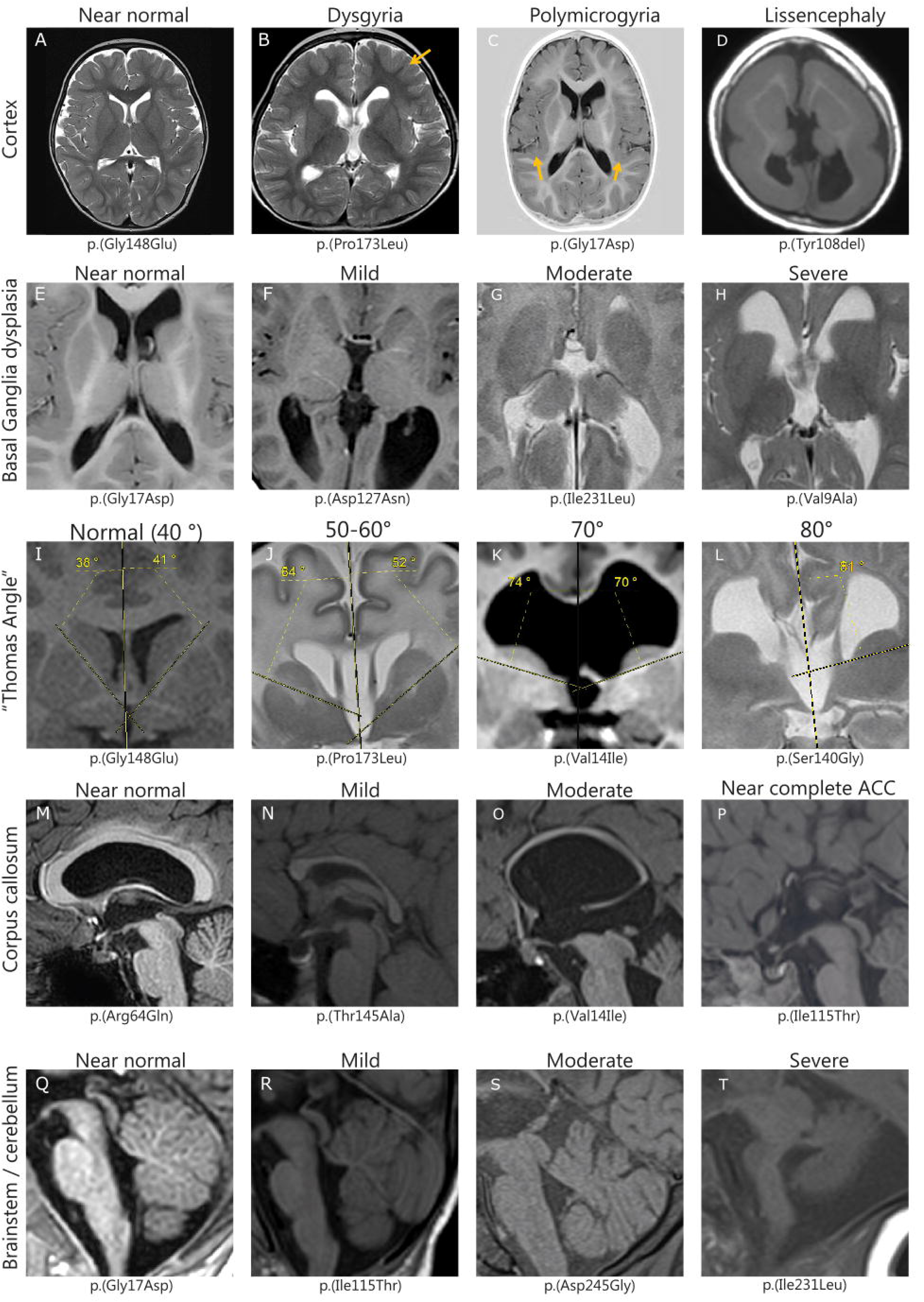
Representative images demonstrating the spectrum of neuroradiological findings in *TUBA1A*-related tubulinopathy. **A-D**: Axial images of: **A**, Individual 12; **B**, Individual 13; **C**, Individual 5; **D**, Individual 7 Orange arrows indicate focal areas of cortical abnormality. **E-H**: Axial images of **E**, Individual 5 **F**, Individual 9; **G**, Individual 17; **H**, Individual 1 **I-L**: Coronal images of **I**, Individual 12; **J**, Individual 16; **J**, Individual 3; **L**, Individual 10 Yellow dotted lines represent planes of measurement **M-P**: Sagittal images of **M**, Individual 6; **N**, Individual 11; **O**, Individual 3; **P**, Individual 8 **Q-T**: Sagittal images of **Q**, Individual 5; **R**, Individual 8; **S**, Individual 18; **T**, Individual 17

Considering the cortex, 5/20 individuals had a normal or near normal appearance. 10/20 demonstrated dysgyria (**Figure 1B**) and 4/20 had polymicrogyria with some simplification of gyral sulcation (**Figure 1C**). Widespread lissencephaly was noted only in **Individual 7** (**Figure 1D**). Despite the paucity of overt cortical abnormalities, all individuals displayed at least one radiographic feature consistent with the diagnosis of a tubulinopathy, with aplasia/hypoplasia of the corpus callosum the most common (17/20). The imaging feature most specific to tubulinopathies, malrotation of the basal ganglia, was also seen in 17/20 individuals. This was clearly apparent in the context of otherwise mild and non-specific features in 6 individuals (**Individuals 1-3, 8, 10 and 23**). The internal capsule angle on coronal imaging could be calculated in 17/20 individuals (**Table** 1 and **Supplemental Table 4**), ranging from 38 degrees to 81 degrees. Average internal capsule angle close to normal (40-45 degrees), suggesting lack of basal ganglial malrotation, was closely related with low cumulative radiological severity score (Supplemental Table 5). Six individuals (**Individuals 6, 11-14 and 20**) represented the mildest neuroradiological abnormalities in the cohort, with a cumulative radiological severity scores between 1 and 5 out of 18.

### Clinical findings

Detailed case-by-case clinical summaries are present in the **Supplementary Text**, with a synopsis in **Table 1** and **Supplemental Table 6.** Global developmental delay was a universal feature. Specifically, median ages of smiling (19.5 weeks, n=10, typically 6 weeks), sitting (1 year 9 months, n=14, typically 6 months) and walking (3 years 7 months, n=16, typically 1 year) were more than three times the typical age of achievement reflecting a severe-profound degree of impairment (**Supplemental Table 7**).

Considering epilepsy, the prevalence of afebrile seizures was not statistically different to that previously reported (60% [12/20] vs 71.2% [37/51] Chi-square p=0.30).^10^ Microcephaly (Occipitofrontal circumference [OFC] <2S.D), both primary and secondary, was significantly less frequent in our cohort than previously reported (59% [13/22] vs 89% [47/53], Chi-square p=0.0035)^10^. Severe feeding problems in infancy were a common feature in our study (9/17), requiring nasogastric or gastrostomy feeding in at least five cases. Ocular findings including optic nerve hypoplasia (3 patients) and abnormalities of eye movements including nystagmus (7 patients) were common in those who underwent eye examination, although in some cases eye examination was normal.

### *in vitro* functional studies

We performed functional assessment of 16 *TUBA1A* variants, categorising them based on the distinctly different functional consequences compared with wild-type *TUBA1A* (**Figure 2**, **Figure 3, Supplemental Figures 2-5, Table 1**)

**Figure 2:**
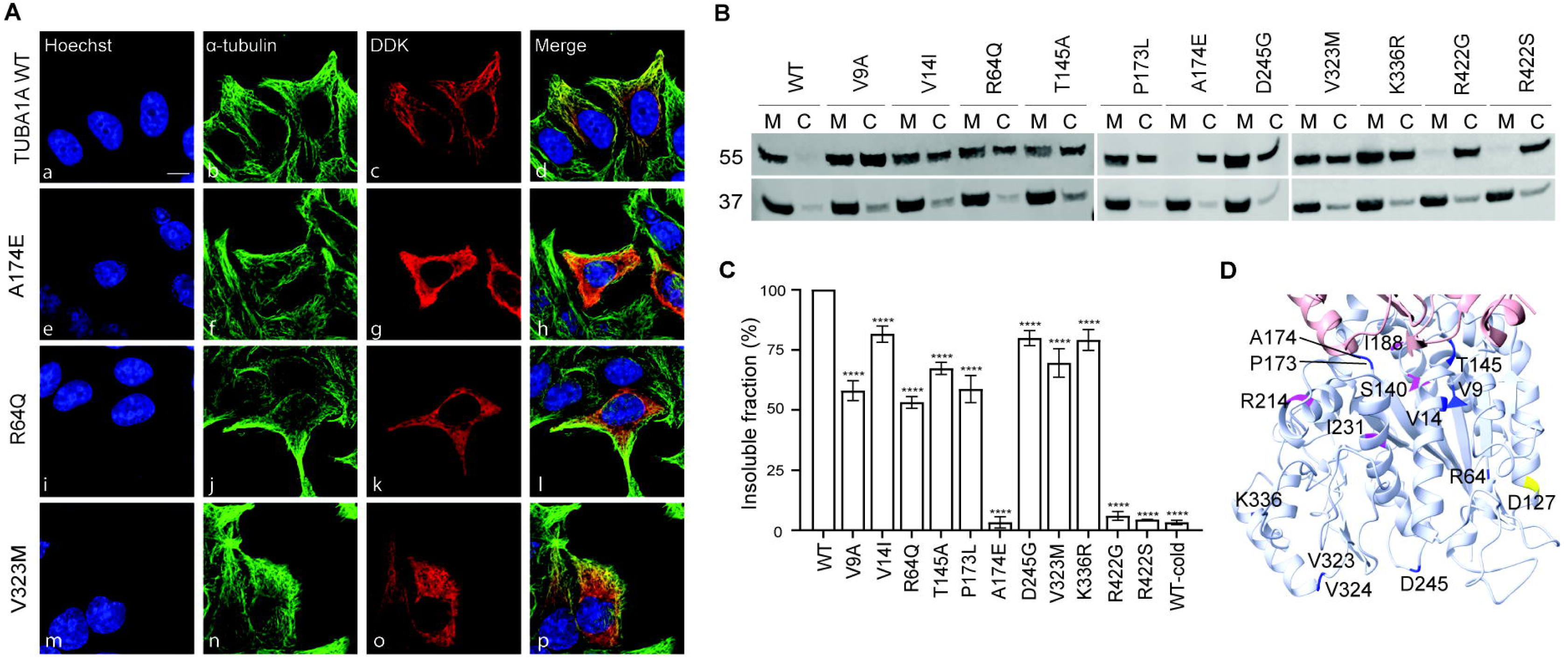
Significant reduction in the physiological incorporation of eleven TUBA1A variants. (**A**) Representative immunofluorescent staining of the endogenous alpha-tubulin (green; panels b, f, j and n) and DDK-tagged transfected tubulin (red; panels c, g, k and o) showed significant reduction in the colocalization of the signals for the variants (panels h, l and p) compared to the TUBA1A wild-type (panel d) demonstrating a varying degree of reduced incorporation of the variants into the microtubules at a physiological level. The p.(Ala174Glu) variant represented an almost absence of incorporation (panel h), the p.(Arg64Gln) variant had slightly higher incorporation (panel l), while the p.(Val323Met) variant had the most incorporation (panel p). (**B-C**) Western blotting to visualise (B) and quantify (C) the amount of transfected protein in the microtubule (M) and cytosolic (C) fraction also confirmed the reduced microtubular incorporation for these variants, compared to the TUBA1A wild-type. Incorporation into the microtubules was measured as the percent of the tagged protein in the insoluble fraction normalised to wild-type as 100% and presented as mean ± SEM (****p<0.001). (**D**) Wild-type TUBA1A showing all variants except p.(Arg422Glu) and p.(Arg422Ser). The variants are colour-coded according to in-vitro findings: decreased level of depolymerisation (pink), decreased level of repolymerisation (yellow), and reduced level of incorporation (blue).

**Figure 3:**
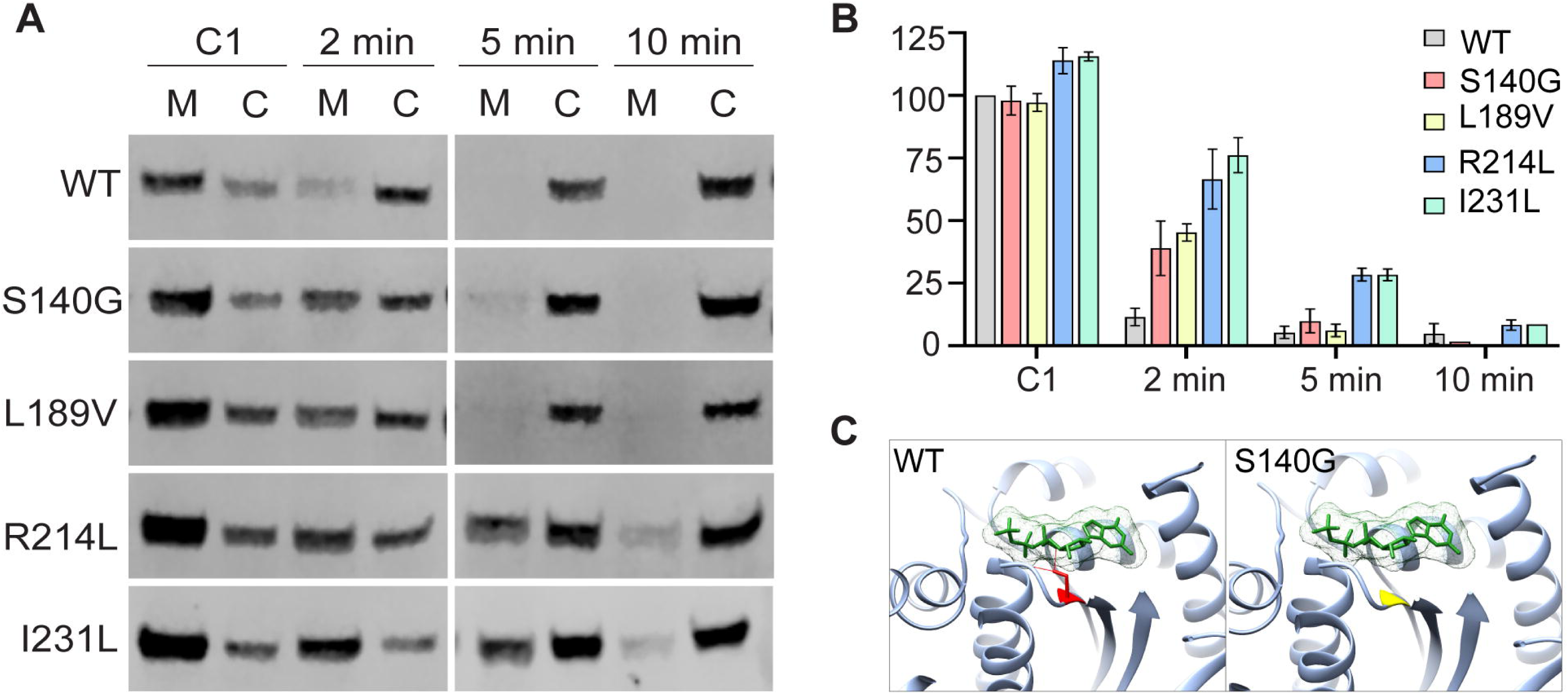
Four TUBA1A variants display slower depolymerisation dynamics compared to the wild-type. (**A, B**) Comparison of the cold-induced depolymerisation dynamics for TUBA1A wild-type and four variants (p.(Ser140Glu), p.(Leu189Val), p.(Arg214Leu) and p.(Ile231Leu)) showed a slower rate of depolymerisation for all four variants. The wild-type completely depolymerised when incubated for 2 minutes at 4°C as demonstrated by the lack of signal in the microtubule fraction (M), while all four variants were significantly located in the microtubules. The p.(Ser140Gly) and p.(Leu189Val) variants completely depolymerised after being at 4°C for 5 minutes. For the variants p.(Arg214Leu) and p.(Ile231Leu), there was a small amount of the transfected tubulin signal in the microtubule fraction (M) at 10 minutes cold incubation, suggesting a significant defect in cold-induced depolymerisation dynamics. Incorporation into the microtubules was measured as the percent of the tagged protein in the insoluble fraction normalised to wild-type as 100% and presented as mean ± SEM (****p<0.001). (**C**) Structural model of wild-type TUBA1A with bound GTP molecule (green) and the p.(Ser140Gly) variant. Left panel shows the hydrogen bonds (red) between the GTP molecule and the S140 residue (red). Right panel shows the disruption of hydrogen bonds at the variant site of the p.(Ser140Gly) variant (yellow) in *TUBA1A* which may compromise the GTP binding and subsequently lead to a reduction in polymerisable tubulin heterodimers.

#### Three TUBA1A variants did not incorporate into the microtubule network

Substituting the alanine at position 174 with glutamate p.(Ala174Glu), and the arginine at position 422 with either glycine or serine p.(Arg422Gly/Arg422Ser) completely abolished the ability of TUBA1A incorporation into the microtubule network. This was observed both by immunofluorescence (**Figure 2A**) through the lack of colocalization between the DDK-tagged variant (red) and endogenous tubulin (green), as well as through an increase of the DDK-tagged variant tubulin in the cytosolic fraction in western blotting (**Figure 2B**). Ala174 is proximal to the GTP-binding pocket, and the substitution of the small, non-polar alanine with a larger, polar glutamate is predicted to reduce the number of hydrogen bond with Ser178, a residue positioned within the ribose binding loop (α/ β: 162-181). This alteration may affect the bonding between Ala174 and GTP (**Supplemental Figure 6**) ^12^.

In addition, Arg422 is located on the external surface in the C-terminal H11-H12 loop (**Supplemental Figure 6**), and the substitution of arginine with Gly422 or Ser422 is predicted to lose hydrogen bonds with D396 and interactions with D392. All associated residues are positioned within the microtubule-associated proteins (MAPs) binding domain^12^, and the loss of interactions among these residues would alter their binding with motor proteins and MAPs^12^.

The studied TUBA1A variants were localised in five main domains (**Figure 2D & Supplemental Figure 6**): interdimer and intradimer interfaces, lateral contact surface, GTP binding region and external MAP binding sites. However, a discernible pattern connecting mutational effects to specific variant locations was not identified.

#### Eight TUBA1A variants showed significantly reduced incorporation into the microtubule network

TUBA1A variants p.(Val9Ala), p.(Val14Ile), p.(Arg64Gln), p.(Thr145Ala), p.(Pro173Leu), p.(Asp245Gly), p.(Val323Met) and p.(Lys336Arg) showed a significantly reduced incorporation into the microtubule network as compared to the *TUBA1A* wild-type (**Figure 2**, **Supplemental Figure 2**). There is some incorporation in the network as seen by the slight colocalization of the transfected variant tubulin with the endogenous tubulin (IF) as well as the presence of some transfected tubulin in the insoluble fraction (WB), albeit at significantly lower levels than that for *TUBA1A* wild-type.

#### The p.(Asp127Asn) variant physiologically incorporated well into the microtubule network, but showed a defect in reincorporation

The p.(Asp127Asn) *TUBA1A* variant showed a similar incorporation into the microtubule network as the wild-type, both by immunofluorescence and western blotting (**Supplemental Figures 3-4**). We further investigated the effect of this variant on microtubule repolymerisation following cold-induced depolymerisation. To do this, microtubules of the transfected cells were allowed to depolymerise at 4°C for 30 minutes and the recovery or reincorporation of tubulin at 37°C for 15 minutes was examined. Wild-type TUBA1A recovered completely within 15 minutes as evidenced by the colocalization (IF) and the abundance of the transfected protein in the insoluble fraction (WB; **Supplemental Figure 3**). The variant showed a significant decrease in the colocalized signal (**Supplemental Figure 3A**) as well as the protein in the insoluble fraction (WB) following recovery when compared to the wild-type (**Supplemental figure 3B-C**).

#### Variants with no discernible change in the microtubule incorporation, but showed a slower rate of depolymerisation

TUBA1A variants p.(Ser140Gly), p.(Leu189Val), p.(Arg214Leu) and p.(Ile231Leu) showed a comparable physiological incorporation to the wild-type, with the variants p.(Arg214Leu) and p.(Ile231Leu) showing similar or even significantly higher incorporation, respectively (**Supplemental Figure 5**). None of these variants demonstrated any defect in repolymerisation following cold-induced depolymerisation. Due to the unique property of variants showing higher incorporation and seemingly no defects, we hypothesised that these variants could be more tightly incorporated into the microtubules than the wild-type equivalent. This would still impact the microtubule dynamics and would be evident if the rate of depolymerisation was investigated. Therefore, the rate of depolymerisation was quantified by incubating the cells for different timepoints (2min, 5min and 10min) at 4°C.

Wild-type TUBA1A completely depolymerised within 2 minutes, as evidenced by a shift of most of the tagged protein into the soluble cytosolic fraction (**Figure 3A**). For all four variants, there was a significant delay in this process as they still had a significant portion of the tagged protein in the insoluble microtubule fraction, suggesting a defect in depolymerisation (**Figure 3B**). The p.(Ser140Gly) and p.(Leu189Val) variants depolymerised completely by 5 minutes, while both p.(Arg214Leu) and p.(Ile231Leu) variants still had a significant amount of tagged tubulin protein remaining in the microtubule fraction. It took 10 minutes for complete depolymerisation of these two variants comparable to the TUBA1A wild-type.

3D modelling analysis of the p.(Ser140Gly) variant showed a loss of interaction with GTP, potentially compromising the ability of the heterodimer to effectively bind to GTP (**Figure 3C**). This could subsequently lead to a reduction in polymerisable tubulin heterodimers. A previous study on mice heterozygous for the p.(Ser140Gly) variant indicated a reduced formation of polymerisable tubulin heterodimers attributing to a compromised GTP binding pocket.^8^

### Clinical-mechanistic associations

We sought to ascertain whether an association exists between the mechanism seen in our functional assays and the clinical or neuroradiological phenotype observed, but this was not definitive for any of the four mechanisms. Whilst instances of polymicrogyria appear more frequently among individuals harbouring variants that exhibit reduced polymerisation (2/4 individuals vs 2/16 individuals) and they had greater severity of intellectual disability (3/4 with severe-profound intellectual disability), the limited number of individuals in this subset precludes a definitive assessment of the significance of these findings. The absence of tubulin incorporation, which might be expected to have the greatest impact on microtubule abundance, did not align with the most severe clinical and neuroradiological manifestations; for example, **Individual 14**, heterozygous for p.(Ala174Glu), presented with a near-normal MRI and only moderate global developmental delay, and **Individuals 6 and 22**, heterozygous for p.(Arg64Gln) and p.(Arg422Gly) respectively, exhibited only mild developmental delay.

## Discussion

We report the first systematically assembled cohort of individuals with pathogenic or likely pathogenic *TUBA1A* variants ascertained independently of their neuroradiological or neuropathological findings complete with their detailed clinical and neuroradiological phenotypes. Our work has revealed an under-recognised phenotypic group in this disorder with characteristic brain findings of a tubulinopathy, but without gross evidence of a malformation of cortical development (MCD). These individuals have a distinct range of, mostly unpublished, variants broadening the genotypic spectrum in this disorder. In each case assessed, *in vitro* defects of tubulin incorporation or depolymerisation could be demonstrated, the latter not previously described as a causative mechanism of *TUBA1A*-related tubulinopathy.

The most significant discovery in our cohort is the low penetrance of overt cortical malformations (5 out of 20 individuals), four of whom had polymicrogyria and only one individual an overt MCD in the form of lissencephaly. In contrast, 10 out of 20 had milder phenotypes including localised (8/20) or diffuse dysgyria (1/20) and 5 out of 20 had normal cortical appearances. These finding are in marked contrast to the collective conclusions of prior publications summarised by Hebebrand *et al*.,^10^ that estimated a penetrance of nearly 99% (76/77 individuals) for overt MCDs. The disparities observed are unlikely to be attributable to benign variants in our cohort, since the novel variants have been robustly assessed according to the ACMG/AMP criteria, with assertions of pathogenicity supported by functional data. Moreover, the presence of specific tubulinopathy-associated neuroradiological findings in the form of basal ganglial malrotation, and additional supportive imaging features including ventriculomegaly, hypo/dysplasia of the corpus callosum, cerebellum and brainstem in most of the cohort, confirms the affected status of individuals and defines a new sub-class of clinical presentation.^19,20^

We propose that a gap existed in previous literature, stemming from an ascertainment bias that favoured the recruitment, testing, and reporting of subjects exhibiting radiological or histopathological features of MCDs,^19,21–24^ thereby overrepresenting these individuals. Applying similar recruitment criteria to our cohort would have excluded a majority of the affected individuals, suggesting that worldwide a significant number of individuals with *TUBA1A*-related tubulinopathy may remain unsuspected, untested and unpublished.

Whilst sporadic reports of *TUBA1A* variants in individuals without MCDs have arisen from other cohorts with unrelated phenotypes (intellectual disability,^11^ autism,^25^ cerebral palsy,^26^ and others^27–30^), no systematic examination of these individuals has occurred. This omission may be attributed to the challenges inherent in conclusively establishing the pathogenicity of novel missense variants in this gene when associated phenotypes diverge from the published literature. Indeed, it is conceivable that many *TUBA1A* variants identified in published studies remain unreported, further compounding the aforementioned evidence gap. We sought to bridge this gap, overcoming the stated challenges by integrating functional studies into a large cohort (in excess of 13,600) with recruitment independent of radiological findings.

A question arising from our research pertains to the penetrance of cortical malformations in *all* individuals with pathogenic variants in *TUBA1A*. This issue, of penetrance in unselected populations, is poised to gain prominence in the coming years as an increasing number of individuals harbouring pathogenic *TUBA1A* variants are expected to be identified through the increasing use of prenatal exome and genome sequencing and possibly through genome sequencing of newborns.^31^ For these individuals, it is likely that our findings provide a better estimate of penetrance for *TUBA1A*-associated phenotypes than the previous literature. However, it is conceivable that our cohort may be depleted of some individuals with MRI findings of overt cortical dysplasia. This group, which tends to be overrepresented in prior literature, may be less likely to be present in DDD having been diagnosed using targeted gene-panel testing prior to recruitment. If this is so, our data might be indicative of the minimum estimated prevalence of cortical malformations among all individuals with *TUBA1A* pathogenic variants.

The absence of MCDs in most individuals in this cohort means that this can no longer be considered as a principal diagnostic neuroradiological sign for *TUBA1A*-related tubulinopathy. In this cohort rotation of the basal ganglia, detected in 17 of the 20 affected individuals, has a much greater sensitivity and likely higher positive-predictive value in tubulinopathies. The anterior commissure was hypoplastic or absent in 17 of the 20 individuals. This finding may be seen in a restricted range of neurogenetic disorders but is not specific to tubulinopathies. Corpus callosal hypo-dysplasia was also observed frequently (17/20), its diagnostic utility is limited due to heterogenous genetic aetiology and resultant low specificity for tubulinopathies.^32^ The prevalence of brain stem dysplasia (14/20) and cerebellar hypoplasia (11/20), both recognised features of tubulinopathies,^33^ was also high in this cohort. Dysmorphology of the basal ganglia is an established feature of tubulinopathies that has been described as the hallmark of these disorders.^33^ This has often been equated with a globular appearance of the basal ganglia nuclei with a fused striatum known as the “absent ALIC sign”. Whilst this represents one end of the spectrum, it does not capture the full range of this feature, which can also be identified when more subtly expressed. The definition and quantification of rotation of the basal ganglia, particularly in relation to the newly established “Internal Capsule Angle”, was a critical aspect of this research, that underlines the significance of this neuroradiological feature as a diagnostic finding in *TUBA1A*-related tubulinopathy.

The only variant in our cohort that has been identified in multiple previous reports is *TUBA1A* p.(Pro173Leu), which we report in **Individual 13** who has severe developmental delay but with only minor neuroradiological abnormalities. This variant has been previously reported in association with polymicrogyria^12^ and as a *de novo* finding in cohorts of individuals with non-syndromic autism and developmental delay,^34,35^ suggesting that the penetrance of overt MCDs for this variant may be low.

The identification of a radiographically unique cohort of individuals with *TUBA1A*-related tubulinopathy with distinctive variants, prompts an examination of whether the clinical phenotype of these individuals also diverges from those typically associated with *TUBA1A*-related tubulinopathy. With regards to microcephaly, despite a permissive definition our cohort demonstrates a lower incidence compared to that reported (76% vs 89%). Whilst this discrepancy is likely attributable to the lower incidence of MCDs in our cohort, since these disorders are frequently associated with microcephaly,^36^ the presence of three individuals in our cohort with near normal neuroimaging and significant microcephaly (**Individual 12, Individual 13, Individual 20**), suggests a degree of independence between these two features in *TUBA1A*-related tubulinopathy.

A notable clinical observation from our cohort is the high frequency of significant feeding difficulties. This does not seem to result in long-term short stature or low body mass index, potentially due to medical interventions, such as nasogastric feeding. While severe feeding challenges in *TUBA1A*-related tubulinopathy have been sporadically reported, they are not typically included as a core feature of the disorder and were not specifically addressed by Hebebrand *et al.*^10^ Our cohort includes six individuals with significant documented feeding difficulties, as many as had been reported in over one hundred previously reported or published individuals. It is also plausible that the neuroradiological focus of many previous studies has led to an incomplete reporting of this and other clinical features (such as epilepsy and developmental delay).

The rich developmental phenotype data in DDD project participants enables detailed analysis of the developmental trajectory associated with *TUBA1A*-related tubulinopathy, especially the age of achievement of developmental milestones (**Supplemental Table 7**). These were universally delayed in our cohort and typically more than three times the typical age of achievement reflecting a severe-profound degree of impairment (**Supplemental Table 7**). Notably, these are conservative estimates of developmental delay since values only contributed to the median if the child’s current age was at least three times the typical age of reaching this milestone. It is likely that in future years these median ages will rise in this cohort.

The mechanism of disease in *TUBA1A*-related tubulinopathy remains a topic of debate.^37–39^ Although there is significant depletion of truncating *TUBA1A* variants in population databases such as gnomAD (LOUEF score = 0.37) and some missense variants do appear to diminish the abundance of mature tubulin, truncating *TUBA1A* variants expected to undergo nonsense-mediated decay are not observed in individuals with *TUBA1A*-related tubulinopathy, making haploinsufficiency an unlikely mechanism.^8,39^ This phenomenon may be partially explained by the genomic architecture of *TUBA1A*, specifically its large final exon which restricts the region where truncating variants would typically be subjected to nonsense-mediated decay to 74 out of 452 codons. Additionally, a resilience of *TUBA1A* against haploinsufficiency has been hypothesised to result from the functional redundancy provided by the array of tubulin paralogues.^40^ Alternative proposed mechanisms in *TUBA1A*-related tubulinopathy implicate perturbations to the secondary, tertiary, and quaternary structural integrity of tubulins. These include an inability of the tubulin dimers to incorporate into microtubules [p.(Leu397Pro)], a dominant negative effect on microtubule stability [p.(Pro263Thr)]^39^ and an accelerated rate of depolymerisation that implies reduced microtubule stability [p.(Cys25Phe), p.(Arg64Trp)].^41^ Certain variants appear to exhibit a spectrum of potential pathogenic mechanisms. Interestingly, some of the most frequently observed variants in radiologically ascertained cohorts [p.(Arg402Cys) & p.(Arg402His)] form functional tubulin heterodimers and assemble into microtubules without difficulty. These variants seem to exert a gain-of-function effect by directly modulating dynein interaction.^42^

Our extensive functional analyses, representing the most comprehensive *in vitro* study of pathogenic *TUBA1A* variants to date, support the notion that there are multiple, variant-specific, molecular mechanisms contributing to the pathogenesis of *TUBA1A*-related tubulinopathies in humans. The majority of the variants identified in this study (11/16) result in reduced tubulin incorporation, presumably due to either a reduction in dimer substrates or a direct failure of incorporation. One variant instead [p.(Asp127Asn)] demonstrated delayed reincorporation. Four variants showed normal incorporation but a reduced rate of depolymerisation, a novel molecular mechanism not previously described in *TUBA1A*-related tubulinopathies. Given the stochastic nature of microtubules, the impact on depolymerisation may be as important as effects on assembly. Whilst increased depolymerisation has been recognised as a mechanism,^41^ a reduced rate of depolymerisation has not been previously observed in *TUBA1A*-related tubulinopathies. This is not routinely sought in cellular assays, meaning it is possible that other variants may exhibit similar *in vitro* behaviours.

Prior to the widespread adoption of gene-agnostic sequencing methodologies for healthcare, most individuals with known *TUBA1A* variants had typically undergone targeted genetic testing for the investigation of a specific phenotype, nearly always an MCD. Under these circumstances the previously published evidence regarding *TUBA1A* was probably adequate for the clinical interpretation of variants identified. However, it is now imperative to reassess our understanding of this disorder, especially regarding penetrance, as widespread adoption of exome and genome sequencing takes place in healthcare. Our research cohort is a more accurate representation of the genotypic and phenotypic spectrum of *TUBA1A*-related tubulinopathies. Insights from this cohort shed light on the nature of *TUBA1A* and tubulinopathies as a group. Characterised by developmental delay and distinctive neuroimaging, yet seldom exhibiting MCDs, these patients represent an emergent phenotypic subgroup. Identification of further similar cohorts will enhance genetic diagnostics, inform appropriate clinical management, and contribute to the scientific comprehension of tubulinopathy mechanisms and potential therapeutic strategies.

## Data Availability

All data produced in the present study are available upon reasonable request to the authors.

## Author Contributions

JF and AD obtained and analysed clinical and genetic data for the whole cohort.

RP, ATH, AVD, RG and MM performed functional studies.

SKC, AVD, ATH and RP interpreted and analysed functional studies data.

KAM, SKC, DK and JGLM generated / interpreted and analysed 3D modelling data.

AGT reviewed the neuroimaging.

SKC, AD, JF, and RP prepared the original draft and reviewed revisions.

NR, KEC. GEJ, MS, AK, TD, KS, WDJ, JR, JAP, ME, ES, CAJ, HVF, SB, RN-E, NG, AJG, SJ, SM, PV and MP performed clinical phenotyping and/or interpreted clinical and genetic results for individual patients.

AD, ES and CAJ supervised the clinical and genetic aspects of the project, contributed to project management and obtained funding for research studies.

SKC designed and supervised functional studies.

All authors reviewed and commented on drafts of the manuscript.

## Acknowledgements

The authors would like to thank all of the individuals and their families who contributed to the DDD project and specifically to those included in this study.

The DDD study presents independent research commissioned by the Health Innovation Challenge Fund [grant number HICF-1009-003], a parallel funding partnership between Wellcome and the Department of Health, and the Wellcome Sanger Institute [grant number WT098051]. The views expressed in this publication are those of the author(s) and not necessarily those of Wellcome or the Department of Health. The study has UK Research Ethics Committee approval (10/H0305/83, granted by the Cambridge South REC, and GEN/284/12 granted by the Republic of Ireland REC).

The research utilised data from the UK Biobank resource carried out under UK Biobank application number 103356. UK Biobank protocols were approved by the National Research Ethics Service Committee. This study was supported by the National Institute for Health and Care Research Exeter Biomedical Research Centre.”

The research team acknowledges the support of the National Institute for Health Research, through the Comprehensive Clinical Research Network. This study makes use of DECIPHER (https://www.deciphergenomics.org), which is funded by Wellcome.

This study was supported by the National Institute for Health and Care Research (NIHR) Exeter Biomedical Research Centre and the NIHR Manchester Biomedical Research Centre (NIHR203308). The views expressed are those of the author(s) and not necessarily those of the NIHR or the Department of Health and Social Care.

JF was supported by as supported by GW4-CAT and Wellcome PhD Training Fellowship for Clinicians 220600/Z/20/Z (https://gw4-cat.ac.uk/, accessed 12/09/2024) and NIHR Exeter Biomedical Research Centre Mid-Career fellowship. We acknowledge support from a University of Leeds Anniversary PhD Studentship (to ME), an MRC project grant (MR/K011154/1 to CAJ) and a Sir Jules Thorn Biomedical Research Award (JTA/09 to ES and CAJ).

## Ethics declarations

UK Research Ethics Committee approval (10/H0305/83, granted by the Cambridge South REC, and GEN/284/12 granted by the Republic of Ireland REC) was granted to the DDD study. All participants consented provided written consent at entry to this study. For all individuals recruited outside of this study consent for research and publication has been obtained from a parent or legal guardian.

The authors have received and archived written patient consent.

## References

1. Barkovich AJ, Guerrini R, Kuzniecky RI, Jackson GD, Dobyns WB. A developmental and genetic classification for malformations of cortical development: update 2012. Brain : a journal of neurology. 2012;135(Pt 5):1348–1369.

2. Guerrini R, Dobyns WB. Malformations of cortical development: clinical features and genetic causes. The Lancet Neurology. 2014;13(7):710–726.

3. Romaniello R, Arrigoni F, Fry AE, et al. Tubulin genes and malformations of cortical development. Eur J Med Genet. 2018;61(12):744–754.

4. Kumar RA, Pilz DT, Babatz TD, et al. TUBA1A mutations cause wide spectrum lissencephaly (smooth brain) and suggest that multiple neuronal migration pathways converge on alpha tubulins. In: Hum Mol Genet. Vol 19.2010:2817–2827.

5. Akhmanova A, Steinmetz MO. Control of microtubule organization and dynamics: two ends in the limelight. Nature Reviews Molecular Cell Biology. 2015;16(12):711–726.

6. Janke C, Magiera MM. The tubulin code and its role in controlling microtubule properties and functions. Nature Reviews Molecular Cell Biology. 2020;21(6):307–326.

7. Wang Z, Sheetz MP. The C-terminus of tubulin increases cytoplasmic dynein and kinesin processivity. Biophys J. 2000;78(4):1955–1964.

8. Keays DA, Tian G, Poirier K, et al. Mutations in alpha-tubulin cause abnormal neuronal migration in mice and lissencephaly in humans. Cell. 2007;128(1):45–57.

9. Poirier K, Keays DA, Francis F, et al. Large spectrum of lissencephaly and pachygyria phenotypes resulting from de novo missense mutations in tubulin alpha 1A (TUBA1A). Hum Mutat. 2007;28(11):1055–1064.

10. Hebebrand M, Hüffmeier U, Trollmann R, et al. The mutational and phenotypic spectrum of TUBA1A-associated tubulinopathy. Orphanet Journal of Rare Diseases. 2019;14(1):38.

11. de Ligt J, Willemsen MH, van Bon BW, et al. Diagnostic exome sequencing in persons with severe intellectual disability. The New England journal of medicine. 2012;367(20):1921–1929.

12. Schröter J, Popp B, Brennenstuhl H, et al. Complementing the phenotypical spectrum of TUBA1A tubulinopathy and its role in early-onset epilepsies. Eur J Hum Genet. 2022;30(3):298–306.

13. The Deciphering Developmental Disorders Study. Large-scale discovery of novel genetic causes of developmental disorders. Nature. 2015;519(7542):223–228.

14. Wright CF, Fitzgerald TW, Jones WD, et al. Genetic diagnosis of developmental disorders in the DDD study: a scalable analysis of genome-wide research data. Lancet. 2015;385(9975):1305–1314.

15. Fourniol FJ, Sindelar CV, Amigues B, et al. Template-free 13-protofilament microtubule-MAP assembly visualized at 8 A resolution. The Journal of cell biology. 2010;191(3):463–470.

16. Cushion TD, Paciorkowski AR, Pilz DT, et al. De novo mutations in the beta-tubulin gene TUBB2A cause simplified gyral patterning and infantile-onset epilepsy. Am J Hum Genet. 2014;94(4):634–641.

17. Smedley D, Smith KR, Martin A, et al. 100,000 Genomes Pilot on Rare-Disease Diagnosis in Health Care - Preliminary Report. The New England journal of medicine. 2021;385(20):1868–1880.

18. Richards S, Aziz N, Bale S, et al. Standards and guidelines for the interpretation of sequence variants: a joint consensus recommendation of the American College of Medical Genetics and Genomics and the Association for Molecular Pathology. Genet Med. 2015;17(5):405–424.

19. Oegema R, Cushion TD, Phelps IG, et al. Recognizable cerebellar dysplasia associated with mutations in multiple tubulin genes. Hum Mol Genet. 2015;24(18):5313–5325.

20. Oegema R, Barakat TS, Wilke M, et al. International consensus recommendations on the diagnostic work-up for malformations of cortical development. Nature Reviews Neurology. 2020;16(11):618–635.

21. Di Donato N, Timms AE, Aldinger KA, et al. Analysis of 17 genes detects mutations in 81% of 811 patients with lissencephaly. Genet Med. 2018;20(11):1354–1364.

22. Bahi-Buisson N, Poirier K, Fourniol F, et al. The wide spectrum of tubulinopathies: what are the key features for the diagnosis? Brain : a journal of neurology. 2014;137(Pt 6):1676–1700.

23. Cushion TD, Dobyns WB, Mullins JG, et al. Overlapping cortical malformations and mutations in TUBB2B and TUBA1A. Brain : a journal of neurology. 2013;136(Pt 2):536–548.

24. Morris-Rosendahl DJ, Najm J, Lachmeijer AM, et al. Refining the phenotype of alpha-1a Tubulin (TUBA1A) mutation in patients with classical lissencephaly. Clinical genetics. 2008;74(5):425–433.

25. Neale BM, Kou Y, Liu L, et al. Patterns and rates of exonic de novo mutations in autism spectrum disorders. Nature. 2012;485(7397):242–245.

26. McMichael G, Bainbridge MN, Haan E, et al. Whole-exome sequencing points to considerable genetic heterogeneity of cerebral palsy. Molecular psychiatry. 2015;20(2):176–182.

27. Kuperberg M, Lev D, Blumkin L, et al. Utility of Whole Exome Sequencing for Genetic Diagnosis of Previously Undiagnosed Pediatric Neurology Patients. Journal of child neurology. 2016;31(14):1534–1539.

28. Posey JE, Rosenfeld JA, James RA, et al. Molecular diagnostic experience of whole-exome sequencing in adult patients. Genet Med. 2016;18(7):678–685.

29. Meng L, Pammi M, Saronwala A, et al. Use of Exome Sequencing for Infants in Intensive Care Units: Ascertainment of Severe Single-Gene Disorders and Effect on Medical Management. JAMA pediatrics. 2017;171(12):e173438.

30. Srivastava S, Cohen JS, Vernon H, et al. Clinical whole exome sequencing in child neurology practice. Ann Neurol. 2014;76(4):473–483.

31. Bick D, Ahmed A, Deen D, et al. Newborn Screening by Genomic Sequencing: Opportunities and Challenges. Int J Neonatal Screen. 2022;8(3).

32. Pânzaru M-C, Popa S, Lupu A, Gavrilovici C, Lupu VV, Gorduza EV. Genetic heterogeneity in corpus callosum agenesis. Frontiers in Genetics. 2022;13 (Review).

33. Gonçalves FG, Freddi TAL, Taranath A, et al. Tubulinopathies. Top Magn Reson Imaging. 2018;27(6):395–408.

34. Fu JM, Satterstrom FK, Peng M, et al. Rare coding variation provides insight into the genetic architecture and phenotypic context of autism. Nature genetics. 2022;54(9):1320–1331.

35. Li J, Wang L, Yu P, et al. Vitamin D-related genes are subjected to significant de novo mutation burdens in autism spectrum disorder. Am J Med Genet B Neuropsychiatr Genet. 2017;174(5):568–577.

36. Lemons JA, Schreiner RL, Gresham EL. Relationship of Brain Weight to Head Circumference in Early Infancy. Human Biology. 1981;53(3):351–354.

37. Bittermann E, Abdelhamed Z, Liegel RP, et al. Differential requirements of tubulin genes in mammalian forebrain development. PLoS genetics. 2019;15(8):e1008243.

38. Hoff KJ, Neumann AJ, Moore JK. The molecular biology of tubulinopathies: Understanding the impact of variants on tubulin structure and microtubule regulation. Frontiers in Cellular Neuroscience. 2022;16 (Review).

39. Tian G, Jaglin XH, Keays DA, Francis F, Chelly J, Cowan NJ. Disease-associated mutations in TUBA1A result in a spectrum of defects in the tubulin folding and heterodimer assembly pathway. Human Molecular Genetics. 2010;19(18):3599–3613.

40. Aiken J, Buscaglia G, Aiken AS, Moore JK, Bates EA. Tubulin mutations in brain development disorders: Why haploinsufficiency does not explain TUBA1A tubulinopathies. Cytoskeleton. 2020;77(3-4):40–54.

41. Yokoi S, Ishihara N, Miya F, et al. TUBA1A mutation can cause a hydranencephaly-like severe form of cortical dysgenesis. In: Sci Rep. Vol 5.2015.

42. Aiken J, Moore JK, Bates EA. TUBA1A mutations identified in lissencephaly patients dominantly disrupt neuronal migration and impair dynein activity. Hum Mol Genet. 2019;28(8):1227–1243.

