## Supplemental for "Multiple and novel molecular mechanisms in *TUBA1A*-related tubulinopathy: insights from deep clinical and neuroradiological phenotyping"

### Supplementary Material

#### Supplemental Table 1: TUBA1A variants identified in this study with population allele frequencies and *in silico* predictions

| Indiv | de novo | NM_006009.4 | Predicted protein outcome | Other reports [PubMed ID] | AlphaMissense (NM_001270399.2) | REVEL | SpliceAI | gnomAD v4.1 | ClinVar (ID) | Decipher ID (this study) |
|---|---|---|---|---|---|---|---|---|---|---|
| 1,2 | Yes | c.26T>C | p.(Val9Ala) | [30744660] [a] | Likely Pathogenic (0.6178) | 0.668 | ≤ 0.2 | Absent | Cf* (625491) | 269057, 276589, 457638 |
| 3,4 | Yes | c.40G>A | p.(Val14Ile) | [30744660] [a] | Likely Benign (0.1788) | 0.208 | ≤ 0.2 | Absent | Absent | 257960 |
| 5 | Yes | c.50G>A | p.(Gly17Asp) | - | Likely Pathogenic (0.9991) | 0.881 | ≤ 0.2 | Absent | P* (1172808) | 550257 |
| 6 | Yes | c.191G>A | p.(Arg64Gln) | - | Likely Pathogenic (0.9803) | 0.866 | ≤ 0.2 | 1 het [f] | LP* (985813) | 296523 |
| 7 | Yes | c.322_324del | p.(Tyr108del) | - | NA | NA | ≤ 0.2 | Absent | Absent | 272013 |
| 8 | Yes | c.344T>C | p.(Ile115Thr) | - | Ambiguous (0.3744) | 0.717 | ≤ 0.2 | Absent | LP/P* (426626) | 284075, 482885 |
| 9 | NK | c.379G>A | p.(Asp127Asn) | 2 papers [b] | Likely Pathogenic (0.9029) | 0.522 | ≤ 0.2 | Absent | LP/P* (427180) | 261467, 332699, 534933, 550069 [e] |
| 10 | Yes | c.418A>G | p.(Ser140Gly) | [30744660] [a] | Likely Pathogenic (0.7386) | 0.82 | ≤ 0.2 | Absent | Absent | 273321 |
| 11 | Yes | c.433A>G | p.(Thr145Ala) | [30744660] [a] | Likely Pathogenic (0.9593) | 0.907 | ≤ 0.2 | Absent | Absent | 283618 |
| 12 | Yes | c.443G>A | p.(Gly148Glu) | - | Likely Pathogenic (0.9999) | 0.276 | ≤ 0.2 | Absent | Absent | 550259 |
| 13 | Yes | c.518C>T | p.(Pro173Leu) | 14 papers [c] | Likely Pathogenic (0.9922) | 0.909 | ≤ 0.2 | Absent | Cf* (625511) | 260435, 379243, 458046 [e], 517452 [e] |
| 14 | NK | c.521C>A | p.(Ala174Glu) | - | Likely Pathogenic (0.9841) | 0.354 | ≤ 0.2 | Absent | Absent | 454597 [e] |
| 15 | Yes | c.565C>G | p.(Leu189Val) | [30744660] [a] | Likely Pathogenic (0.98) | 0.784 | ≤ 0.2 | Absent | Absent | 286470 |
| 16 | Yes | c.641G>T | p.(Arg214Leu) | [34716235] | Likely Pathogenic (0.713) | 0.678 | ≤ 0.2 | Absent | LP/P** (432708) | 304960 |
| 17 | Yes | c.691A>C | p.(Ile231Leu) | [30744660] [a] | Likely Benign (0.2273) | 0.412 | ≤ 0.2 | Absent | LP (3344362) | 279083 |
| 18 | Yes | c.734A>G | p.(Asp245Gly) | [30744660] [a] | Likely Pathogenic (0.9318) | 0.736 | ≤ 0.2 | Absent | Absent | 285013 |
| 19,20 | Yes/NK | c.967G>A | p.(Val323Met) | [35017693] | Likely Pathogenic (0.9796) | 0.681 | ≤ 0.2 | Absent | Cf* (1206718) | 307453[1], 380794, 407731, 491231 [e] |
| 21 | Yes | c.1007A>G | p.(Lys336Arg) | 2 papers [d] | Ambiguous (0.4672) | 0.833 | ≤ 0.2 | Absent | Absent | 285646 |
| 22 | Yes | c.1264C>G | p.(Arg422Gly) | - | Likely Pathogenic (0.9978) | 0.899 | ≤ 0.2 | Absent | Absent | 305931 [e] |
| 23 | Yes | c.1264C>A | p.(Arg422Ser) | [35017693] | Likely Pathogenic (0.9997) | 0.859 | ≤ 0.2 | Absent | Absent | 307140 |

Data as of 14/02/2025 ClinVar's four-star rating system represents the "Review Status" of each submission: **/* reflects number of submitters, data submitted and conflicts.

**Notes**: [a] Paper refers to the individuals in this paper (deposited in Decipher prior to publication); [b] PubMed IDs: 32005694, 33528536; [c] PubMed IDs: 22495306, 25363768, 28407358, 28714951, 30744660, 31785789, 31133750, 34946966, 34011629, 33726816, 35982160, 35017693, 35982159, 36801247; [d] PubMed IDs: 30744660, 38444904 [a];

[e] Decipher record without open-access sharing, which may not be visible to all users. [f] present in one individual in UKBiobank who has a phenotype compatible with a tubulinopathy.

**Abbreviations**: Indiv Individual, Cf: Conflicting, LP: Likely pathogenic, NK; not known, P: Pathogenic

#### Supplemental Table 2: ACMG/AMP variant classification of TUBA1A variants

| Indiv | de novo | NM_006009.4 | Predicted protein outcome | PS2 | PS3 | PS4 [a] | PM1 | PM2 | PM5 | PP2 | PP3 | Classification |
|---|---|---|---|---|---|---|---|---|---|---|---|---|
| 1,2 | Yes | c.26T>C | p.(Val9Ala) | Strong | Supporting | Moderate [Cv,D] | Moderate [c] | Moderate | - | Supporting | - | Pathogenic |
| 3,4 | Yes | c.40G>A | p.(Val14Ile) | Strong | - | - | Moderate [c] | Moderate | - | Supporting | - | Likely pathogenic |
| 5 | Yes | c.50G>A | p.(Gly17Asp) | Strong | - | Supporting [Cv] | - | Moderate | - | Supporting | Supporting | Likely pathogenic |
| 6 | Yes | c.191G>A | p.(Arg64Gln) | Strong | Supporting | - | - | Moderate | Moderate [d] | Supporting | Supporting | Pathogenic |
| 7 | Yes | c.322_324del | p.(Tyr108del) | Strong | - | - | - | Moderate | - | - | - | Likely pathogenic |
| 8 | Yes | c.344T>C | p.(Ile115Thr) | Strong | - | Supporting [Cv] | - | Moderate | - | Supporting | - | Likely pathogenic |
| 9 | NK | c.379G>A | p.(Asp127Asn) | - | Supporting | Supporting [Cv] | Moderate [b] | Moderate | Supporting [e] | Supporting | - | Likely pathogenic |
| 10 | Yes | c.418A>G | p.(Ser140Gly) | Strong | Supporting | - | Moderate [c] | Moderate | - | Supporting | Supporting | Pathogenic |
| 11 | Yes | c.433A>G | p.(Thr145Ala) | Strong | Supporting | - | Moderate [c] | Moderate | - | Supporting | Supporting | Likely pathogenic |
| 12 | Yes | c.443G>A | p.(Gly148Glu) | Strong | Supporting | - | - | Moderate | Supporting [f] | Supporting | - | Likely pathogenic |
| 13 | Yes | c.518C>T | p.(Pro173Leu) | Strong | Supporting | Moderate [Cv,D] | Moderate [c] | Moderate | - | Supporting | Supporting | Pathogenic |
| 14 | NK | c. 521C>A | p.(Ala174Glu) | - | Supporting | Supporting [D] | Moderate [c] | Moderate | Moderate [g] | Supporting | - | Likely pathogenic |
| 15 | Yes | c.565C>G | p.(Leu189Val) | Strong | Supporting | - | - | Moderate | - | Supporting | Supporting | Likely pathogenic |
| 16 | Yes | c.641G>T | p.(Arg214Leu) | Strong | Supporting | Supporting | Moderate [b] | Moderate | Moderate [h] | Supporting | - | Pathogenic |
| 17 | Yes | c.691A>C | p.(Ile231Leu) | Strong | Supporting | Supporting [Cv] | - | Moderate | - | Supporting | - | Likely pathogenic |
| 18 | Yes | c.734A>G | p.(Asp245Gly) | Strong | Supporting | - | - | Moderate | - | Supporting | Supporting | Likely pathogenic |
| 19,20 | Yes / NK | c. 967G>A | p.(Val323Met) | Strong | Supporting | Moderate [Cv,D] | - | Moderate | - | Supporting | - | Pathogenic |
| 21 | Yes | c.1007A>G | p.(Lys336Arg) | Strong | Supporting | Supporting | - | Moderate | - | Supporting | Supporting | Pathogenic |
| 22 | Yes | c.1264C>G | p.(Arg422Gly) | Strong | Supporting | - | Moderate [b] | Moderate | Moderate | Supporting | Supporting | Pathogenic |
| 23 | Yes | c.1264C>A | p.(Arg422Ser) | Strong | Supporting | Supporting | Moderate [b] | Moderate | Moderate | Supporting | Supporting | Pathogenic |

Guidelines applied as per ACGS Best Practice Guidelines for Variant Classification in Rare Disease 2024 (Durkie *et al.*)

**Notes:** [a] In some cases insufficient details were given regarding affected individuals to use in classification; [b] Local enrichment of pathogenic missense variation; [c] GTP binding site (see Supplemental Figure 6); [d] Arg64Trp, PubMed IDs: 26493046, 36658419; [e] Asp127Glu, PubMed ID: 29907476 [f] Gly148Arg, PubMed ID: 36672771; [g] Ala174Val, PubMed IDs: 31833200, 35017693, 32978145, 33528536; [h] Arg214His, PubMed IDs: 24860126, 26130693, 28677066, 29158550, 30744660, 32570172, 38502138 Arg214Cys, PubMed ID: 33726816 [Cv] ClinVar case/s; [D] Decipher case/s

#### Supplemental Table 3: Variants in genes other than TUBA1A identified through the DDD project that could not be otherwise excluded at the time of testing.

| Individual(s) | Gene | Variant | gnomAD v4.1.0 | Inheritance |
|---|---|---|---|---|
| **1** | *CHD8* ENST00000646647.2: | c.2372C>T; p.(Pro791Leu) | 46 heterozygous | Inherited from unaffected father |
| | *KMT2D* ENST00000301067.12 | c.1277T>C; p.(Leu426Pro) | 15 heterozygous | Inherited from unaffected father |
| | *FBN2* ENST00000508053.5 | c.8467C>A; p.(Pro2823Thr) | 17 heterozygous | Inherited from unaffected father |
| **9** | *NEXMIF* ENST00000055682.12 | c.2849A>T; p.(Tyr950Phe) | 114 heterozygous | Inh unknown |
| **11** | *BPTF* ENST00000306378.11 | c.4079_4081del p.Lys1360_Pro1361delinsThr | 62 heterozygous | Inherited from unaffected mother |
| **20** | *PIEZO2* ENST00000503781.7 | c.8228T>C; p.(Met2743Thr) | 1 heterozygous | Inheritance unknown |
| | *DSPP* ENST00000651931.1 | c.762T>A; p.(Asp254Glu) | Absent | Inheritance unknown |
| | *COL4A1* ENST00000375820.10 | c.4831G>A; p.(Ala1611Thr) | 1 heterozygous | Inheritance unknown |
| | *LHX4* ENST00000561113.1 | c.163G>A; p.(Gly55Arg) | Absent | Inheritance unknown |
| **21** | *COL9A1* ENST00000357250.11 | c.1070G>A; p.(Arg357His) | 420 heterozygous | Inherited from unaffected father |
| | | c.2585A>C; p.(Asp862Ala) | 2409 heterozygous | Inherited from unaffected mother |
| | *ABCD4* ENST00000555904.1 | c.163C>A; p.(Pro55Thr) | Absent | Inherited from unaffected mother |
| | | c.463G>T; p.(Asp155Tyr) | 80 heterozygous | Inherited from unaffected father |
| **22** | *OFD1* ENST00000340096.11 | c.2048C>T; p.(Thr683Ile) | Absent | Inheritance unknown |
| | *MAP3K1* ENST00000399503.4 | c.38G>C; p.(Gly13Ala) | 3 heterozygous | Inheritance unknown |
| **2,3,6,7,8, 10, 13, 15, 16, 17, 18, 19, 23** | No variants | | | |
| **4** | Diagnosed using targeted dideoxy (Sanger) sequencing | | | |
| **5, 12, 14** | Not known | | | |

#### Supplemental Table 4: Detailed neuroradiological findings in each reported case

| Ind | NM_006009.4 | Cortex | Basal ganglia | Internal capsule angle | Corpus Callosum | Ant commissure | Cerebellum | Brainstem | Other |
|---|---|---|---|---|---|---|---|---|---|
| **1** | c.26T>C p.(Val9Ala) | Bilateral frontal dysgyria | Abnormal fusion - severe | R - 57 L - 66 | Hypogenesis / hypoplasia | Absent | Inferior vermis hypoplasia | Mild R medulla flattening, thin brainstem sagittal | Asymmetric ventricular dilatation |
| **2** | c.26T>C p.(Val9Ala) | Bilateral frontal dysgyria | Abnormal fusion - severe | T1 coronal imaging not available | Thin | Absent | Mild vermis hypoplasia | Mild pontine hypoplasia | Dysmorphic enlarged frontal horns |
| **3** | c.40G>A p.(Val14Ile) | Bilateral frontal dysgyria | Very flat ALICs - severe | R - 74 L - 70 | Thin | Hypoplastic | Abnormal superior folial pattern, vermis hypoplasia | R>L pons, flat pontomedullary junction | Absent septum pellucidum |
| **4** | c.40G>A p.(Val14Ile) | No MRI | - | - | - | - | - | - | - |
| **5** | c.50G>A p.(Gly17Asp) | Bilateral perisylvian polymicrogyria & parallel frontal sulcation | Minor left-sided hypoplasia | R - 53 L - 54 | Hypoplastic posterior body/splenium | Hypoplastic | Minimal vermian hypoplasia | Narrow midbrain isthmus | Nil |
| **6** | c.191G>A p.(Arg64Gln) | Near normal | Right side mildly abnormally rotated | R - 59 L - 37 | Bulky but fully formed | Hypoplastic | Normal | L>R pons & medulla, homolateral asymmetry, thin medullary isthmus | Asymmetric ventricles |
| **7** | c.322_324del p.(Tyr108del) | Lissencephaly throughout | Completely fused | No ALIC formed | Marked hypoplasia | Absent | Vermian hypoplasia | Pontine hypoplasia | Severe microcephaly |
| **8** | c.344T>C p.(Ile115Thr) | Bilateral frontal dysgyria | Very abnormal L>R | R - 77 L - 77 | Almost absent | Hypoplastic | Mild vermis hypoplasia | Minor right pontine flattening/hypoplasia | Abnormal orientation cerebellar folia |
| **9** | c.379G>A p.(Asp127Asn) | Near normal | Mild left-sided hypoplasia / rotation | R - 53 L - 55 | Very hypoplastic (genu only) | Absent | Normal | Dorsal L pontine tegmental hump | Asymmetric ventricles related to CC dysgenesis |
| **10** | c.418A>G p.(Ser140Gly) | Bilateral frontal dysgyria | Abnormal fusion - severe | R - *, L - 81 | Absent | Absent | Vermis hypoplasia, cortex dysplasia | Very dysplastic, R>L medulla, L>R pons, contralateral asymmetry | Dilated ventricles, thalamic fusion |
| **11** | c.433A>G p.(Thr145Ala) | Near normal | Normal | R - 43 L - 45 | Thin | Absent | Normal | L>R pons (mild) | enlarged ventricles, vertical orientation frontal horns |
| **12** | c.443G>A p.(Gly148Glu) | Near normal | Normal | R - 38 L- 41 | Exaggerated isthmus only | Normal | Normal | Normal | |

| Ind | NM_006009.4 | Cortex | Basal ganglia | Internal capsule angle | Corpus Callosum | Ant commissure | Cerebellum | Brainstem | Other |
|---|---|---|---|---|---|---|---|---|---|
| **13** | c.518C>T p.(Pro173Leu) | Mild dysgyria | Minor right putaminal hypoplasia only | R - 47 L - 45 | Normal | Hypoplastic | Normal | Normal excluding narrow midbrain isthmus | Lateral/3rd ventriculomegaly |
| **14** | c. 521C>A p.(Ala174Glu) | Bilateral minimal frontal dysgyria | Normal | R - 45 L - 41 | Normal | Normal | Normal | Normal | Nil |
| **15** | c.565C>G p.(Leu189Val) | Bilateral perisylvian polymicrogyria, frontal simplified gyral pattern | Abnormal fusion - severe | R - 63 L - 71 | Shortened/ hypoplastic | Absent | Abnormal superior folial orientation, vermis hypodysplasia | R>L medulla, flat pontomedullary junction | Absent septum pellucidum |
| **16** | c.641G>T p.(Arg214Leu) | Mild bilateral frontal dysgyria | Horizontal ALICs, mild | R - 64 L - 52 | Rostrum hypogenesis | Absent | Dysplasia - vermis + hemispheres | Flat L medulla (v mild) | Megalencephaly, tortuous vessels persistent cavum SP. Scaphocephaly |
| **17** | c.691A>C p.(Ile231Leu) | Perisylvian polymicrogyria, Bifrontal polymicrogyria | Abnormal - no ALIC but symmetrical, moderate | Unable to measure | Severe hypoplasia | Absent | Vermis hypoplasia, cortex dysplasia | Dysplastic ++, kinked | Absent septum pellucidum |
| **18** | c.734A>G p.(Asp245Gly) | Bifrontal dysgyria and perisylvian polymicrogyria | Abnormal - R ALIC, L flat, severe | R - 60 L - 58 | Hypogenesis | Hypoplastic | Dysplasia - vermis + hemispheres | contralateral asymmetry, L>R pons, R>L medulla, flat pontomedullary junction | Microcephaly |
| **19** | c. 967G>A p.(Val323Met) | Diffuse dysgyria | mildly dysmorphic | R - 46 L - 46 | Hypoplastic | Hypoplastic | Normal | hypoplastic pons with flattening on the right | - |
| **20** | c. 967G>A p.(Val323Met) | Normal | Normal | R - 45 L - 44 | Elongated and flattened | Normal | Normal | Normal | Microcephaly |
| **21** | c.1007A>G p.(Lys336Arg) | No MRI | - | - | - | - | - | - | - |
| **22** | c.1264C>G p.(Arg422Gly) | No MRI | - | - | - | - | - | - | - |
| **23** | c.1264C>A p.(Arg422Ser) | Bilateral posterior short gyrus and bifrontal dysgyria | Abnormal fusion - severe | R-45 L - 70 | Short | Hypoplastic | mild inf vermian hypoplasia only | Mild L medullary < R, R pons < L | Nil |

***** Could not be measured. **Abbreviations:** ALIC, Anterior limb of the internal capsule; CC, Corpus callosum; Cavum SP, cavum septum pellucidum; L, left; R, right
d) sagittal T1 showing hypoplastic rostrum, genu and anterior body of corpus callosum

#### Supplemental Table 5: Radiological severity score calculation

| Indiv | Cortex | Basal Ganglia | CC | Anterior commissure | Cerebellum | Brainstem | Severity score (total 18) |
|---|---|---|---|---|---|---|---|
| **1** | 1 | 3 | 2 | 3 | 1 | 1 | 11 (moderate) |
| **2** | 1 | 3 | 1 | 3 | 1 | 1 | 10 (moderate) |
| **3** | 1 | 3 | 1 | 2 | 1 | 1 | 9 (moderate) |
| **4** | | | | | | | |
| **5** | 2 | 1 | 2 | 2 | 0 | 0 | 7 (moderate) |
| **6** | 0 | 1 | 0 | 2 | 0 | 1 | 4 (mild) |
| **7** | 3 | 3 | 3 | 3 | 2 | 1 | 15 (severe) |
| **8** | 1 | 3 | 3 | 2 | 1 | 1 | 11 (moderate) |
| **9** | 0 | 1 | 3 | 3 | 0 | 1 | 8 (moderate) |
| **10** | 1 | 3 | 3 | 3 | 2 | 3 | 15 (severe) |
| **11** | 0 | 0 | 1 | 3 | 0 | 1 | 5 (mild) |
| **12** | 0 | 0 | 1 | 0 | 0 | 0 | 1 (mild) |
| **13** | 0 | 1 | 0 | 3 | 0 | 1 | 5 (mild) |
| **14** | 1 | 0 | 0 | 0 | 0 | 0 | 1 (mild) |
| **15** | 2 | 3 | 1 | 3 | 1 | 1 | 11 (moderate) |
| **16** | 1 | 1 | 1 | 3 | 2 | 0 | 8 (moderate) |
| **17** | 2 | 2 | 3 | 3 | 2 | 3 | 15 (severe) |
| **18** | 2 | 3 | 2 | 2 | 2 | 2 | 13 (severe) |
| **19** | 1 | 1 | 2 | 2 | 0 | 1 | 7 (moderate) |
| **20** | 0 | 0 | 1 | 0 | 0 | 0 | 1 (mild) |
| **21** | | | | | | | |
| **22** | | | | | | | |
| **23** | 1 | 3 | 1 | 2 | 1 | 1 | 9 (moderate) |

Each neuroradiological feature was graded (0, normal; 1, mild changes; 2, moderate changes; 3, severe changes).
The anterior commissure was also graded (0, normal; 1, hypoplastic; 3, absent).
Summed scores were separated into mild (0-6), moderate (7-12) and severe (13-18) categories.

#### Supplemental Table 6: Detailed clinical findings in each reported case

| Individual | 1 | 2 | 3 | 4 | 5 | 6 | 7 | 8 | 9 | 10 | 11 | 12 |
|---|---|---|---|---|---|---|---|---|---|---|---|---|
| **TUBA1A variant [a]**<br>**First report[1]** | c.26T>C<br>p.(Val9Ala)<br>VCV625491 | c.26T>C<br>p.(Val9Ala)<br>VCV625491 | c.40G>A<br>p.(Val14Ile)<br>This family[2] | c.40G>A<br>p.(Val14Ile)<br>This family[2] | c.50G>A<br>p.(Gly17Asp)<br>VCV1172808 | c.191G>A<br>p.(Arg64Gln)<br>VCV985813 | c.322_324del<br>p.(Tyr108del)<br>This family | c.344T>C<br>p.(Ile115Thr)<br>VCV426626 | c.379G>A<br>p.(Asp127Asn)<br>32005694 | c.418A>G<br>p.(Ser140Gly)<br>This family[2] | c.433A>G<br>p.(Thr145Ala)<br>This family[2] | c.443G>A<br>p.(Gly148Glu)<br>This family |
| ***Inheritance*** | *de novo* | *de novo* | *de novo*[†] | *de novo*[†] | *de novo* | *de novo* | *de novo* | *de novo* | unknown | *de novo* | *de novo* | *de novo* |
| **Growth [ S.D.] –** *Age and Sex Removed to comply with MedRx policies* | | | | | | | | | | | | |
| *Height* | -0.6 | NK | NK | NK | -1.6 | +0.5 | +0.4 | -1.1 | +1.4 | NK | +0.5 | NK |
| *Weight* | -1.1 | -0.7 [c] | -2.8 [c] | -2.8 [c] | -1.0 | +0.1 | -0.1 | -2.6 | +3.3 | -1.9 | +0.6 [c] | NK |
| *Birth OFC* | NK | NK | -1.6 | -1.6 | NK | NK | -3.3 | -2.4 | NK | Microcephaly | NK | NK |
| *OFC* | -2.9 | -3.4 | +0.4 | -0.4 | -0.6 | -1.2 | -6.7 | -6.6 | +1.0 | -7.5 | -0.6 | Microcephaly |
| **Neurology** | | | | | | | | | | | | |
| *Tone* | ↓& brisk reflex | NK | ↓ | ↓ | ↑ | NK | ↓ | NK | NK | Dystonic movements | Normal | Lower limb dystonia |
| *Seizures* | No | No | Yes | Yes | No | No | Yes | Yes | Yes - Febrile | Yes | No | NK |
| *Type* | - | - | Focal / GTCS | Focal / GTCS | - | - | Myoclonic | NK | NK | - | - | NK |
| *Onset* | - | - | 9y | 7y | - | - | NK | NK | NK | - | - | NK |
| *Hearing loss* | ✓ | No | NK | NK | NK | NK | NK | NK | NK | | No | NK |
| *Eye* | ✓ON hypoplasia | No | ✓OM Apraxia | ✓OM apraxia | ✓Abnormal movement | - | NK | NK | NK | ✓CVI, myopia | Normal | ✓Abnormal movements |
| **Development** | | | | | | | | | | | | |
| *Delay / ID* | Severe | Severe | Severe | Severe | Moderate | Mild | Severe | Severe | Moderate | Profound | Mild/Mod | Mild |
| *Behaviours* | NK | NK | NK | NK | Noise sensitivity | NK | NK | NK | NK | NK | ASD | NK |
| **Other** | | | | | | | | | | | | |
| *Feeding difficulties* | No | No | No | No | No | No | ✓NG | No | Yes | Yes - gastrostomy | No | NK |
| *Other* | - | - | - | Scoliosis | Cranio-synostosis | Single palmar crease | hirsutism constipation | NK | Obesity High palate | Scoliosis | Pelvic kidney | Right sided ptosis |

| **Individual**<br>**TUBA1A variant** [a]<br>**First report**[1]<br>***Inheritance*** | 13<br>c.518C>T<br>p.(Pro173Leu)<br>35017693<br>*de novo* | 14<br>c. 521C>A<br>p.(Ala174Glu)<br>35017693<br>unknown | 15<br>c.565C>G<br>p.(Leu189Val)<br>This family[2]<br>*de novo* | 16<br>c.641G>T<br>p.(Arg214Leu)<br>VCV432708<br>*de novo* | 17<br>c.691A>C<br>p.(Ile231Leu)<br>This family[2]<br>*de novo* | 18<br>c.734A>G<br>p.(Asp245Gly)<br>This family[2]<br>*de novo* | 19<br>c. 967G>A<br>p.(Val323Met)<br>35017693<br>*de novo* | 20<br>c.967G>A<br>p.(Val323Met)<br>35017693<br>unknown | 21<br>c.1007A>G<br>p.(Lys336Arg)<br>This family[2]<br>*de novo* | 22<br>c.1264C>G<br>p.(Arg422Gly)<br>This family<br>unknown | 23<br>c.1264C>A<br>p.(Arg422Ser)<br>This family[3]<br>*de novo* |
|---|---|---|---|---|---|---|---|---|---|---|---|
| **Growth [ S.D.] –** *Age and Sex Removed to comply with MedRx policies* | | | | | | | | | | | |
| *Height* | NK | NK | NK | -3.7 | -3.8 | NK | +0.3 | +1.1 | NK | -0.3 | NK |
| *Weight* | -0.3 | NK | -0.9 [c] | -1.9 | -1.2 | -5.1 | NK | +2.2 | NK | -1.5 | NK |
| *Birth OFC* | NK | NK | Normal | NK | +0.4 | NK | NK | -2.8 | NK | NK | NK |
| *OFC* | -3.2 | NK | -2.7 | -0.9 [c] | -5.1 | Microcephaly | +0.7 | -2.1 | Microcephaly | -1.8 | -3.2 |
| **Neurology** | | | | | | | | | | | |
| *Tone* | NK | NK | NK | ↓ | ↓ | ↓ | ↓ | NK | ↓ | NK | ↑ |
| *Seizures* | Yes - | Yes | No | No | Yes | Yes | NK | No | Yes | NK | Yes |
| *Type* | GTCS / myoclonic | Absences | - | - | GTCS / myoclonic | GTCS / myoclonic | NK | - | GTCS / myoclonic | NK | NK |
| *Onset* | | 9m | - | - | 1d | Infancy | NK | - | Infancy | NK | NK |
| *Hearing loss* | NK | NK | NK | NK | NK | No | No | No | Normal | NK | NK |
| *Eye* | NK | NK | Normal | ✓Congenital nystagmus | ✓CVI & ON hypoplasia | Normal | hypermetropia | ✓ON cupping | NK | NK | NK |
| **Development** | | | | | | | | | | | |
| *Delay / ID* | Severe | Moderate | Moderate | Profound | Severe | Severe | Moderate | Moderate | Severe | Mild | Severe |
| *Behaviours* | Self-injurious | Autistic traits | | | | | | | | | |
| **Other** | | | | | | | | | | | |
| *Feeding difficulties* | Yes - NG | NK | Yes | Yes - NG | Yes - PEG & fundoplication | Yes - As infant | NK | NK | Yes - PEG | NK | NK |
| *Other* | | | | Familial Polydactyly Crypt-orchidism | Ant. pituitary hypoplasia adrenal insufficiency | Thoracic scoliosis | Aganglionic megacolon | Eczema | Aganglionic megacolon died, seizure | | |

↑ increased ↓ decreased † Not present in parents, inheritance suggests germline mosaicism [a] NM_006009.4; [b] Age at measurement; [c] measurements taken at a different age (see supplementary text).
**Abbreviations:** ant, anterior**;** CVI, Cortical visual impairment; d, day(s); F, female; GTCS, Generalised tonic-clinic seizures; ID, intellectual disability, (s); M, male; Mod, moderate; NG, nasogastric (tube feeding); *NK, not known;* OM, Oculomotor; ON Optic Nerve; PEG, Parenteral gastrostomy; S.D, standard deviations;. Height, weight, BMI, and OFC Z-scores were calculated using a Microsoft Excel add-in to access growth references based on the LMS (Lambda Mu Sigma) method using a reference European population (https://www.healthforallchildren.com/ , accessed September 3, 2021).

#### Supplemental Table 7: Ages of achievement of developmental milestones

*These have been removed to comply with BioRx policies*
*They are expected to be published in full in the final version*

**Supplemental Figure 1: Comparative genograms of the distribution of previously reported variants and those reported in this publication.**

Larger circles represent multiple reports of the same variant. Figure produced using https://github.com/joiningdata/lollipops

Reported in this study

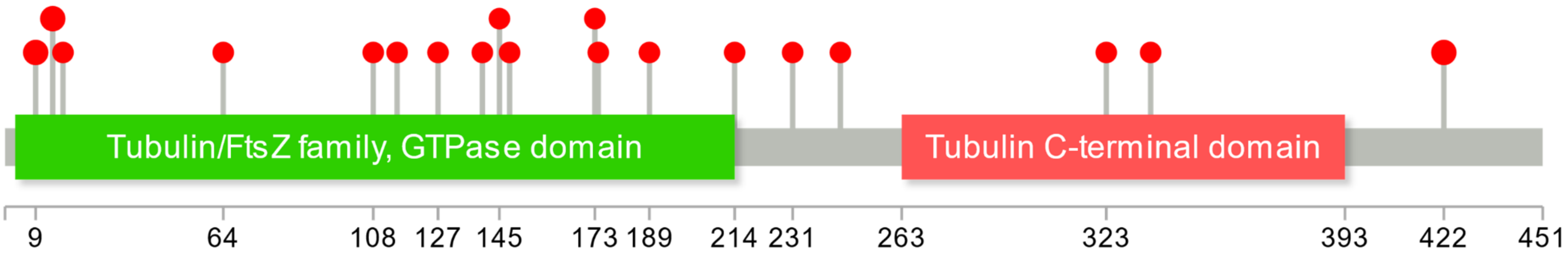


Reported by Hebebrand *et al* 2019

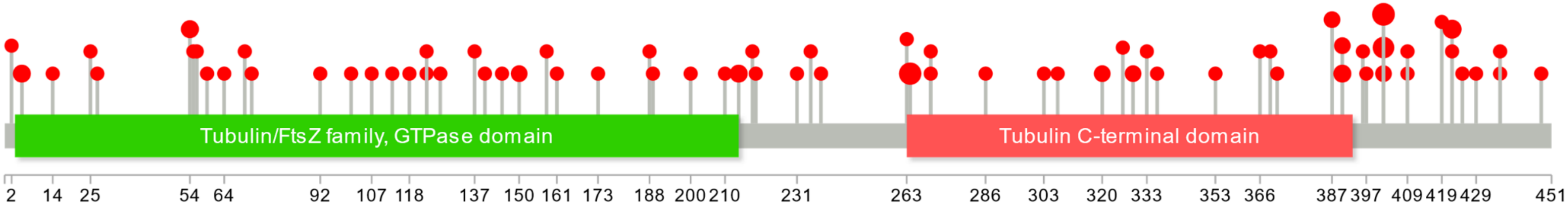

#### Supplemental Figure 2: Reduced incorporation of eight TUBA1A variants

Immunofluorescence staining of eight TUBA1A variants showed a reduced incorporation into the microtubules seen as the colocalization (merge) of the endogenous α-tubulin (green) and DDK-tagged transfected tubulin (red).

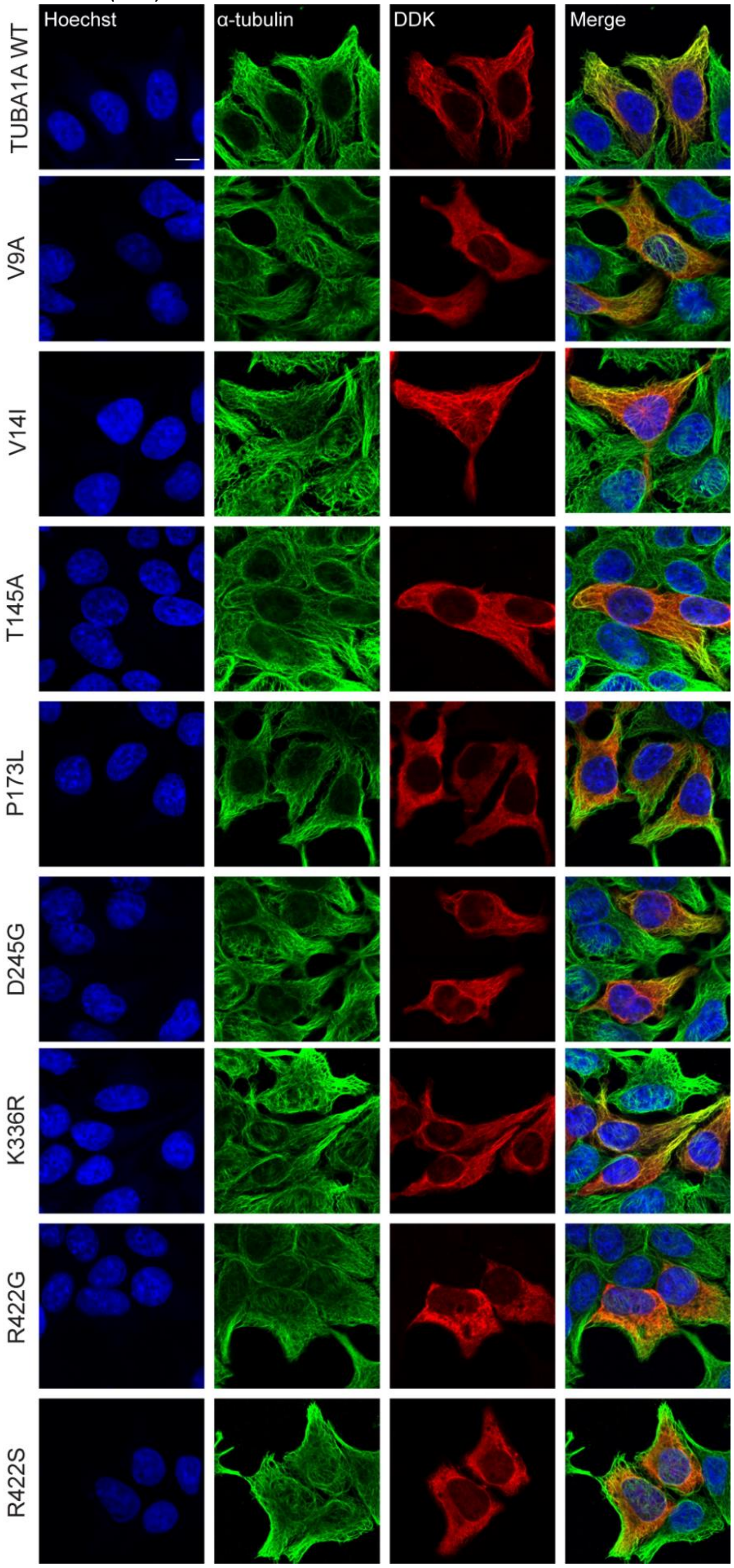

**Supplemental Figure 3: Reduced reincorporation following cold-induced depolymerisation for the D127N variant.**

**A**) Immunofluorescent staining of the endogenous α-tubulin (green; panels b, f, j and n) and DDK-tagged transfected tubulin (red; panels c, g, k and o) showed similar physiological incorporation (C1) of the p.(Asp127Asn) variant (panel h) compared to the TUBA1A wild-type (panel d). Following a 30-minute depolymerisation in cold (4°C) and a 15-minute recovery at 37°C (C3), the p.(Asp127Asn) variant shows significantly reduced reincorporation as seen by the reduced colocalization (panel p) as compared to the TUBA1A wild-type (panel l).

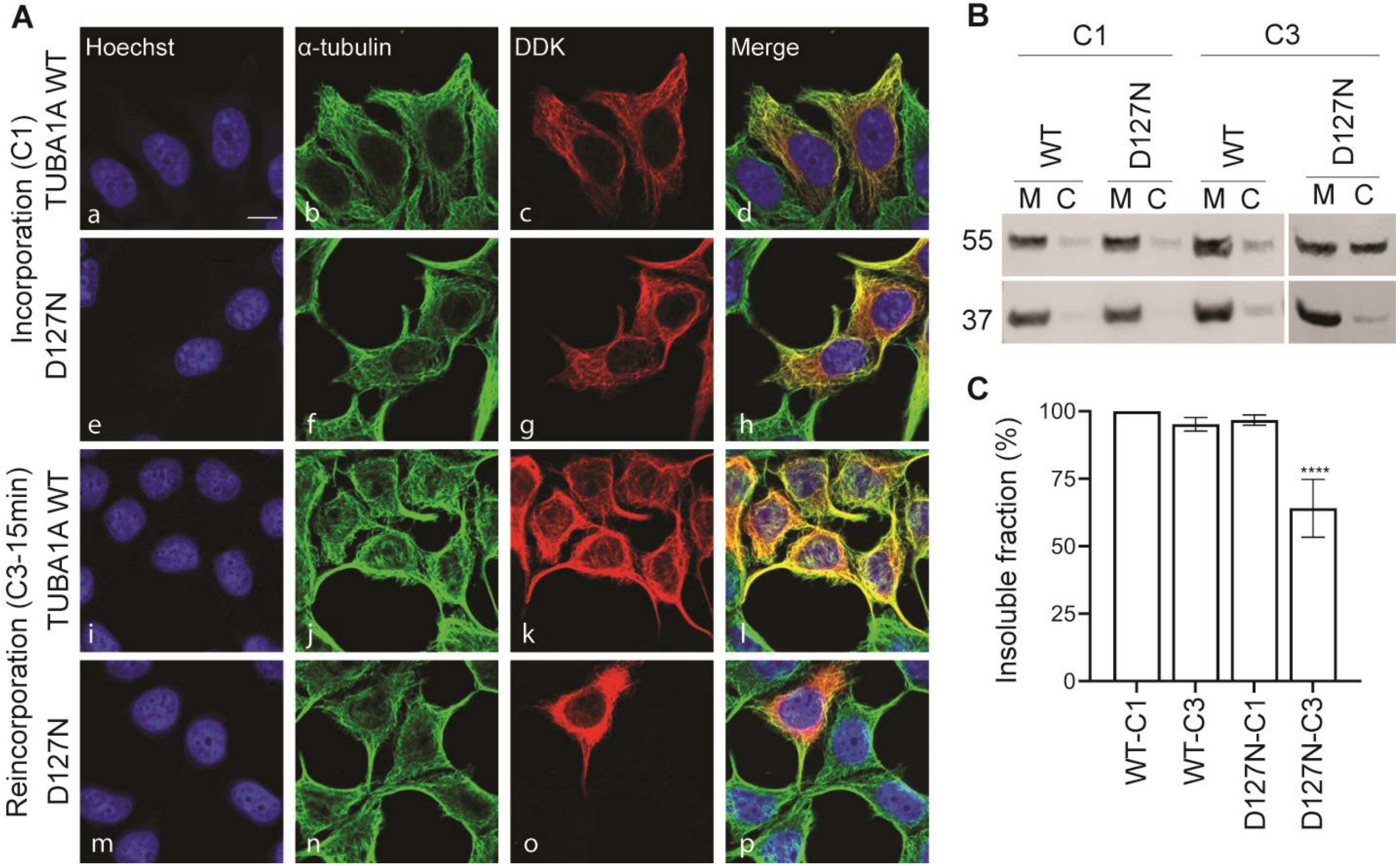


**B**) Reduced reincorporation dynamics for the p.(Asp127Asn) variant are confirmed by western blotting, initially showing a similar physiological incorporation (C1) for the variant compared to the wild-type and a reduced incorporation following a cold-induced depolymerisation (C3), quantified in (C). Incorporation into the microtubules was measured as the percent of the tagged protein in the insoluble fraction normalized to wild-type as 100% and presented as mean ± SEM (****$p<0.001$).

**Supplemental Figure 4: Dynamics of the D127N variant**

The p.(Asp127Asn) variant undergoes complete depolymerisation, similar to the wild-type, when incubated at 4°C for 30 minutes. This is visualised by a complete lack of fibrous staining for both the intrinsic (green) and transfected (red) tubulin, which are assumed to be completely cytosolic. (B) While there is a delay in its reincorporation (see figure 2); the variant completely recovers and incorporates similar to the wild-type when incubated at 37°C for 30 minutes following cold-induced depolymerisation.

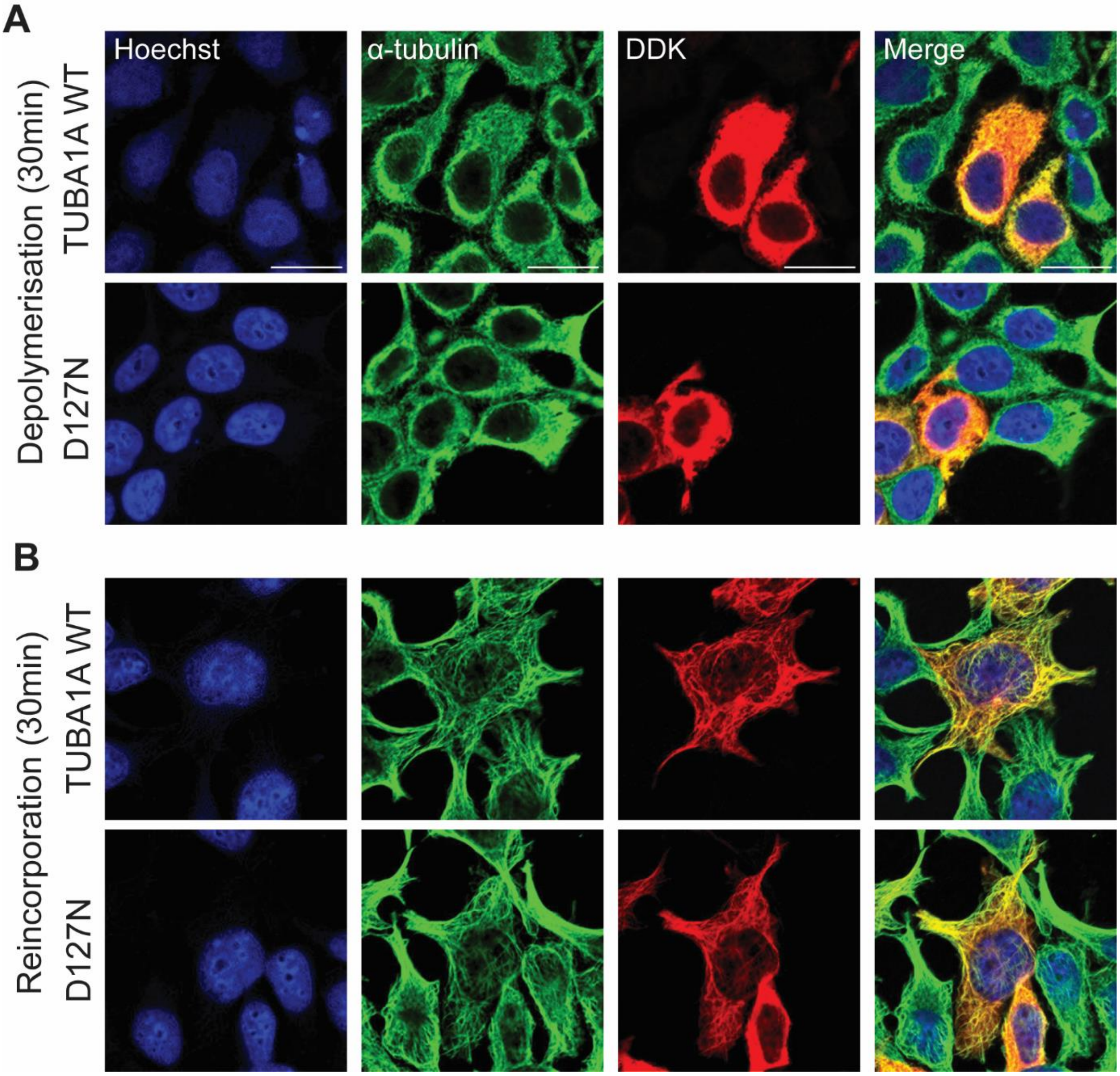

#### Supplemental Figure 5: Four variants do not show any change in gross depolymerisation, and reincorporation dynamics compare to the TUBA1A wild type.

**A**) The variants p.(Ser140Gly), p.(Leu189Val), p.(Arg214Leu) and p.(Ile231Leu) depolymerised completely, as the TUBA1A wild-type, when incubated at 4°C for 30 minutes (cold), visualised through the loss of fibre-like staining for both the intrinsic (green) and transfected (red) proteins
**B**) All four variants also reincorporated at a similar rate to the wild type, when incubated at 37°C for 15 minutes (C3).

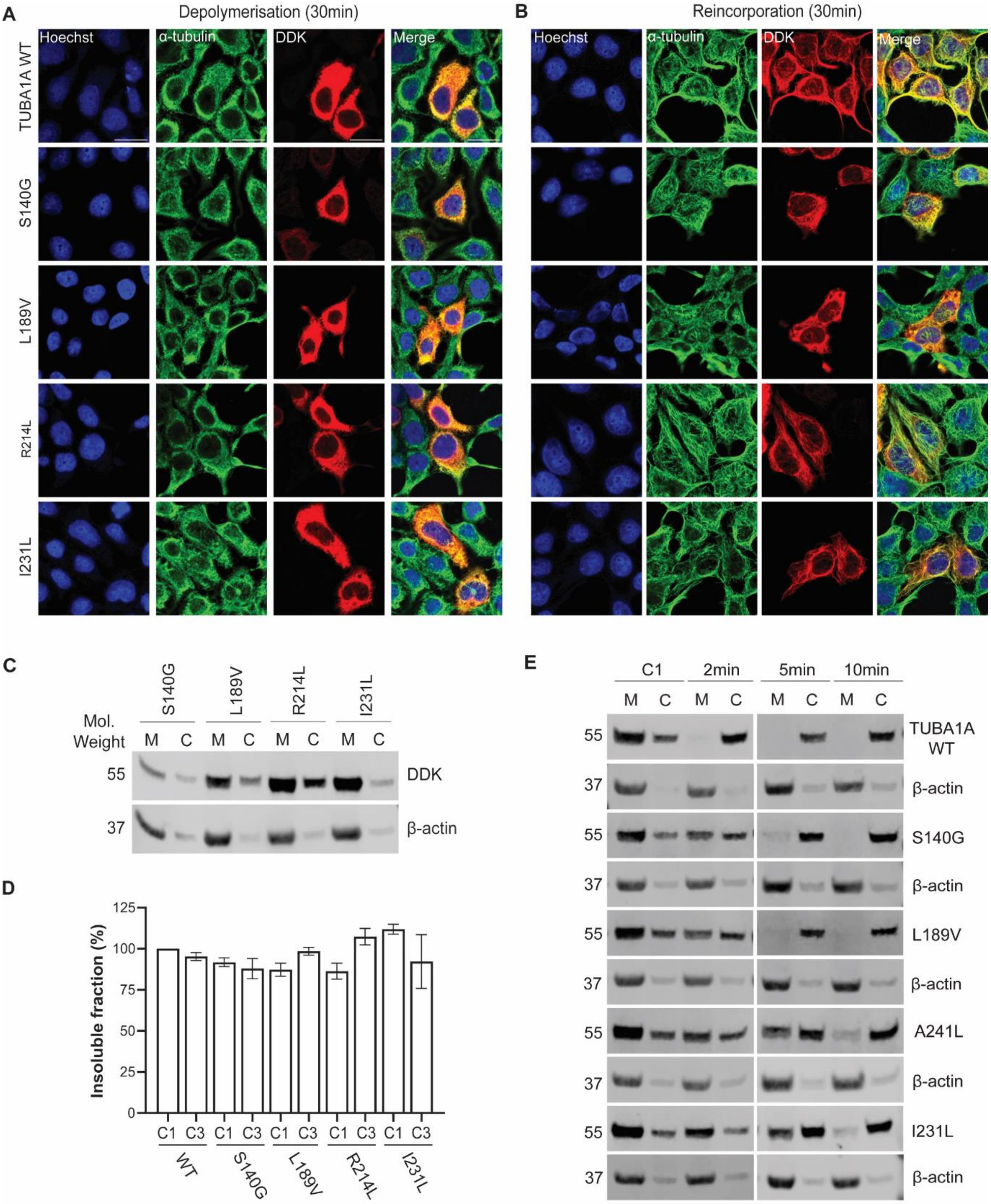


**C,D**) Western blotting showed a similar, if not a higher (trending to, not significant) reincorporation (C3) for the variants compared to the wild-type and their own physiological (C1) incorporation.
**E**) Extended western blot for figure 3, showing the housekeeping protein β-actin (37 kDa).

#### Supplemental Figure 6: Protein modelling demonstrates impact of missense variants in TUBA1A

**A)** Wild-type TUBA1A (α-tubulin shown in blue, and β-tubulin shown in pink) with bound GTP molecule (green) showing all variants except p.(Arg422Glu) and p.(Arg422Ser). The variants are colour-coded according to in-vitro findings: decreased level of depolymerisation (pink), decreased level of repolymerisation (yellow), and reduced level of incorporation (blue).
**B)** Structural model of the α/β-tubulin heterodimer complex with bound GTP molecule (green) and six TUBA1A variants located within the GTP binding region colour-coded according to in-vitro findings. Blue (n=5) indicates reduced level of incorporation and pink (n=1) indicates decreased level of depolymerisation.
**C)** Structural model of the wild-type α-tubulin (blue) heterodimer complex with bound GTP molecule (green) and p.(Arg422Gly)/ p.(Arg422Ser) variants. Both p.(Arg422Gly) and p.(Arg422Ser) variants lead to the loss of all interactions with D396 and D392 residues which are located within the H11 helix, positioned at the interface between α-tubulin and MAPs.
**D)** Structural model of the wild-type α/β-tubulin heterodimer complex and the p.(Ala174Glu) variant. The left panel shows the two hydrogen bonds (in red) formed between the A174 residue and S179. The right panel indicates that substituting alanine with glutamic acid does not lead to notable structural alterations despite the reduced incorporation observed in in-vitro results.

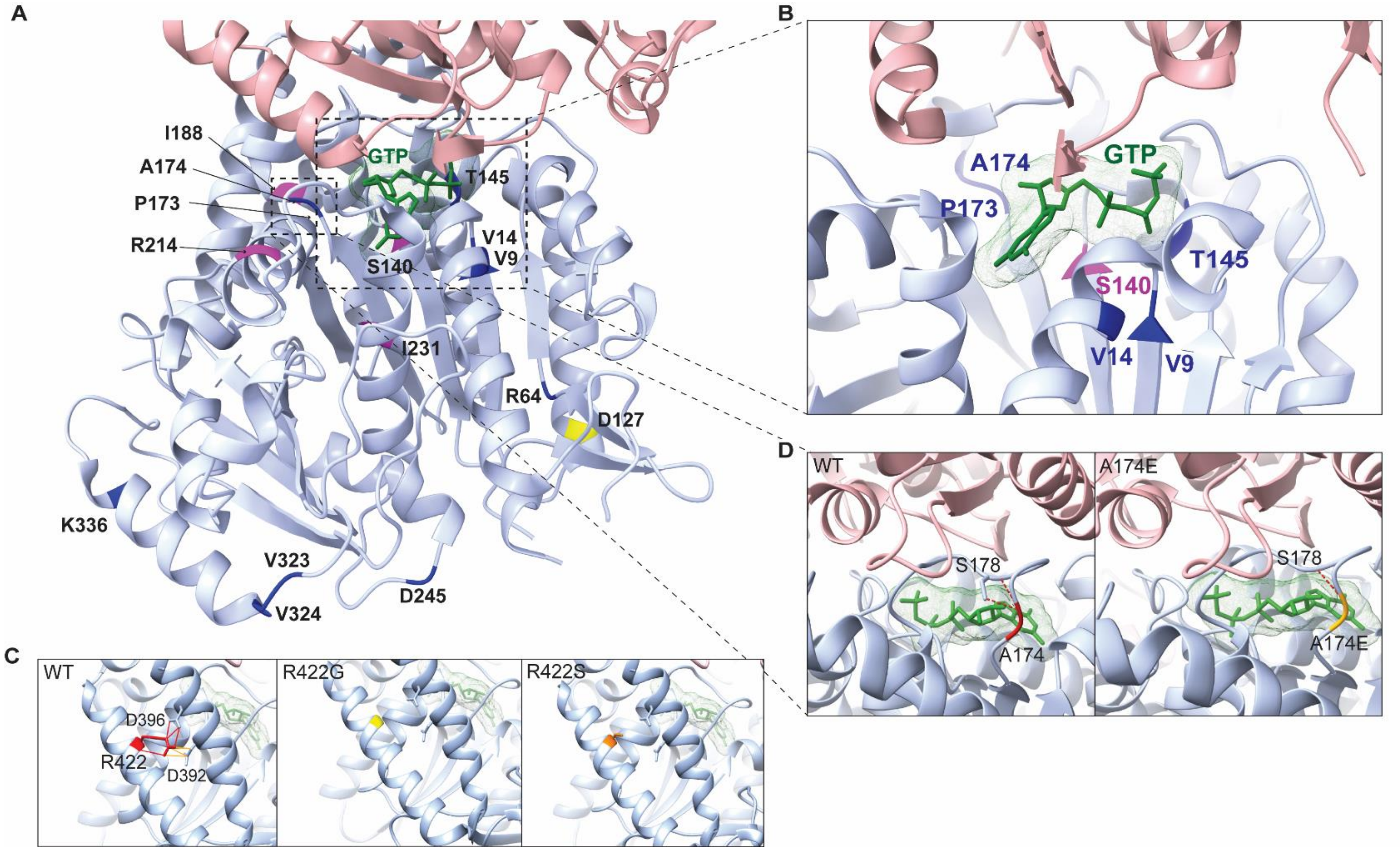

#### Supplemental text: clinical and neuroradiological reports

*These have been removed to comply with BioRx policies*
*They are expected to be published in full in the final version*
